# Modelling treatment effects for gonorrhoea

**DOI:** 10.1101/2023.07.03.23292181

**Authors:** Pavithra Jayasundara, David. G. Regan, Philip Kuchel, James. G. Wood

## Abstract

*Neisseria gonorrhoeae* (NG) bacteria have evolved resistance to many of the antibiotics that have been used successfully to treat gonorrhoea infection. To gain a better understanding of potential treatment options for gonorrhoea, we extend a previously developed within-host mathematical model to integrate treatment dynamics by accounting for key pharmacokinetic (PK) and pharmacodynamic (PD) features. This extended model was used to investigate different treatment regimens for two potential treatment options, namely, monotreatment with gepotidacin, and dual treatment with gentamicin and azithromycin. The simulated treatment success rates aligned well with the, albeit limited, clinical trial data that are available. The simulation results indicated that antibiotic treatment failure is associated with failure to successfully clear intracellular NG (NG residing within epithelial cells and neutrophils) and that extracellular PK indices alone cannot differentiate between treatment success or failure. We found that the index defined by the ratio of area under the curve to minimum inhibitory concentration (AUC/MIC) index > 150h, evaluated using intracellular gepotidacin concentration, successfully distinguished between treatment success and failure. For the dual treatment regimen, AUC/MIC index > 140h evaluated using the simulated single drug concentration, representing the combined effect of gentamicin and azithromycin with the Loewe additivity concept, successfully differentiated between treatment success and failure. However, we found this PK threshold associated with dual treatment to be less informative than in the gepotidacin monotreatment case as a majority of samples below this threshold still resulted in infection clearance. Although previous experimental results on the killing of intracellular NG are scarce, our findings draw attention to the importance of further experiments on antibiotic killing of intracellular NG. This will be useful for testing putative new anti-gonorrhoea antibiotics.

**Author Summary:** Gonorrhoea is a sexually transmitted infection caused by bacteria of the species *Neisseria gonorrhoeae* (NG). Although gonorrhoea can be easily treated using antibiotics, due to the propensity of NG to acquire resistance to antimicrobials, available treatment options have greatly diminished and most of the antibiotics used to treat infection in the past are now removed from treatment recommendations. As clinical trials have limitations in terms of expense, duration and ethical constraints they are not ideal for optimising doses, regimens and drug combinations. In this case, simulations through within-host mathematical models are useful in determining the effective dosing regimens and to explore intracellular treatment effects for which there is little experimental evidence. Our simulations identified the importance of treating intracellular NG (NG residing within neutrophils and epithelial cells) and the importance of considering intracellular pharmacokinetic indices when differentiating treatment success and failure. With the use of this model, we can simulate a range of different treatment regimens and drug combinations to assess their effectiveness at various values of the minimum inhibitory concentration which can potentially be used to guide future clinical trial design.

## Introduction

Gonorrhoea is a sexually transmitted infection caused by bacteria of the species *Neisseria gonorrhoeae* (NG). Since the beginning of the antibiotic era, NG has progressively developed resistance to the classes of drugs used to treat gonorrhoea, and current treatments are now under threat with few alternatives of proven safety and efficacy [1, 2]. Drug resistant NG has become a major public health concern [3, 4] and the development of new treatment options and prophylactic vaccines is seen as increasingly important in population control of gonorrhoea.

In clinical trial settings both gepotidacin (GEP) [5, 6] and gentamicin (GEN) + azithromycin (AZM) dual treatment [7-9] have shown potential for treating urethral NG infection. Gepotidacin is a novel triazaacenaphthylene bacterial type II topoisomerase inhibitor while azithromycin is a macrolide and gentamicin is an aminoglycoside. Both macrolides and aminoglycosides work by disrupting bacterial protein synthesis by inhibiting ribosome functionality [10]. Clinical trials report much higher treatment effectiveness using dual therapy with gentamicin + azithromycin (100% cure rate [8]) than with gentamicin monotherapy (68-98% cure rate [11]), but similar effectiveness to using azithromycin monotherapy (99.2% cure rate [12]). By comparing the minimum inhibitory concentration (MIC) of azithromycin and gentamicin on NG strains under monotherapy and dual therapy, the *in vitro* study by Xu et al. [13] has shown that when used in combination, gentamicin can decrease the progression of the development of azithromycin resistance. This combination therapy is recommended as an alternative treatment for patients who cannot be treated with the recommended treatment ceftriaxone, due to infection with ceftriaxone resistant strains [13], allergy or unavailability of ceftriaxone.

Although clinical trials are considered the gold standard for evaluating the safety and effectiveness of new drugs, they have limitations in terms of expense, duration and ethical constraints, which compromise their utility for optimising doses, regimens and drug combinations [14]. In this case, simulations through compartment pharmacokinetic (PK)/ pharmacodynamic (PD) models such as those used in the study by Chisholm et al. [15] are useful in determining effective dosing regimens. In the context of NG, a within-host mechanistic model has the potential to explore intracellular treatment effects for which there is little experimental evidence. In our previous work on within-host modelling of natural NG infection [16], we observed that intracellular survival and replication of NG appears to be a key factor in prolonging untreated infection. Therefore, it is of interest to consider how treatment resolves infection while accounting for intracellular NG states.

In this context intracellular PK/PD effects appear likely to be essential in guiding the design of treatment regimens. However, while experimental studies of extracellular PK/PD effects for NG infection (e.g., [17, 18]) have been conducted, we were unable to find any studies that explored intracellular PK/PD effects in the context of NG infection.

In this study, we extend the mathematical model of male urethral NG infection developed in Jayasundara et al. [16] to include antibiotic treatment effects. Here we also investigate the impact of intracellular NG in determining MIC for treatments evaluated in recent trials as future options: different dosing strategies using monotreatment with gepotidacin (GEP) and dual treatment with gentamicin (GEN) + azithromycin (AZM). Finally, we analyse intracellular PK/PD dynamics for these regimens and determine intracellular drug concentration levels required for treatment success.

## Materials and Methods

### Mathematical model of antibiotic treatment

In Jayasundara et al. [16], we developed a deterministic compartmental within-host transmission model to describe untreated symptomatic male urethral infection with NG. In that model, four NG states (unattached NG (*B*), NG attached to epithelial cells (B_a_), NG internalised within epithelial cells (B_i_) and NG surviving within polymorphonuclear leukocytes (PMN) (B_S_)) and the innate immune response mediated by PMN are used to describe the infection process. In this study, we extend this model to include treatment effects by applying PK/PD principles. Treatment effects are incorporated in both extracellular (*B* and B_a_) and intracellular NG states (B_i_ and B_S_) using drug-specific Hill functions [19], with differing concentrations of drug in the extracellular and intracellular environments. The Hill function parameters are estimated using the NG growth data reported in the *in vitro* time-kill experiments for gentamicin and azithromycin conducted by Foerster et al. [19] and, for gepotidacin, in the study by Farrell et al. [20]. Further details are described in Appendix S1 Section S1. The dual treatment effects of gentamicin and azithromycin are modelled using the concept of Loewe additivity as these drugs have similar targets and mechanisms of action [7, 21]. When modelling gepotidacin concentration, we adopt a one-compartment model [22] as has been applied by So et al. [23] where we assume that drug concentration declines exponentially on a time-scale determined by the half-life of the drug. However, for gentamicin [24, 25] and azithromycin [26, 27], we adopt a two-compartment model to account for more complex intracellular drug distribution and accumulation. Model specific parameter values are given in Table 1, with the treatment model described in greater detail in the Appendix S1 and the parameters describing untreated infection described in detail in Jayasundara et al. [16]. Fig. 1 provides a schematic illustration of the natural infection model with the added treatment effects.

**Fig 1:**
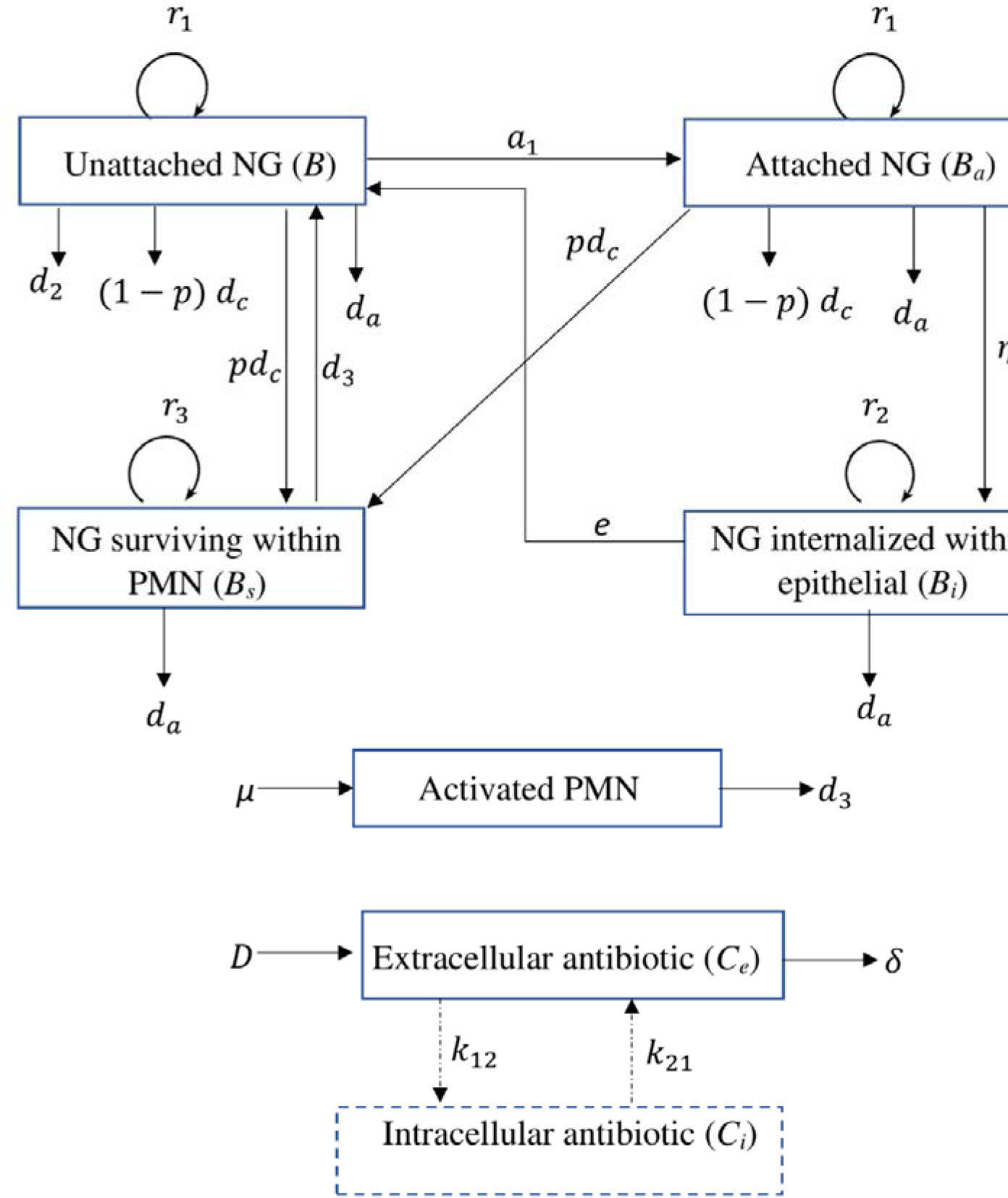
Schematic illustration of the within-host NG infection model including antibiotic treatment. Arrows indicate transitions between model states (boxes). Antibiotic- and PMN-mediated killing of NG are denoted as and, respectively (for killing by PMN see Jayasundara et al. [16]). Explicit intracellular antibiotic compartments are included for gentamicin and azithromycin (see Section ‘Mathematical model of antibiotic treatment’), with transitions between extra and intracellular drug concentrations (dashed lines) applying only for these two drugs.

**Table 1:**
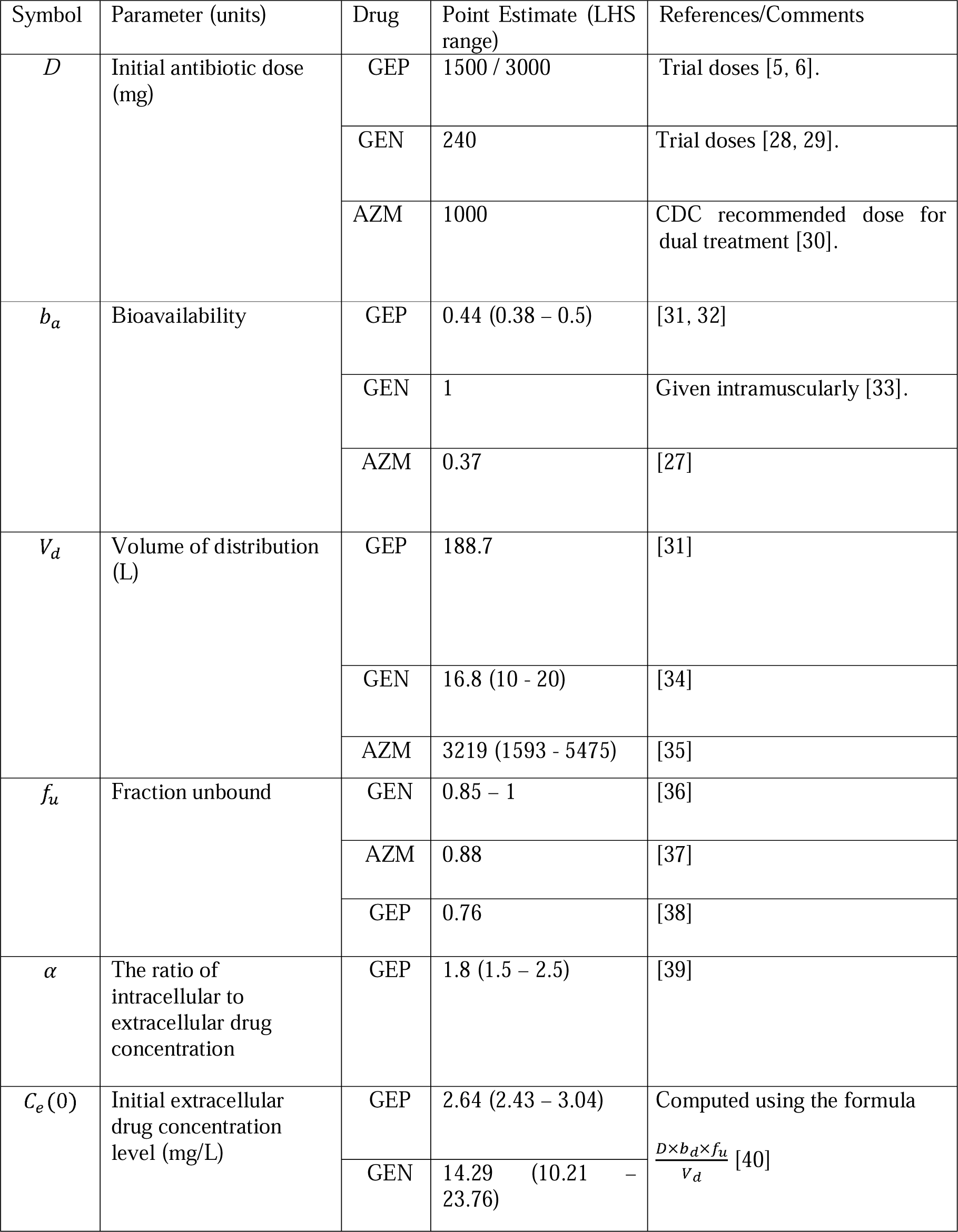
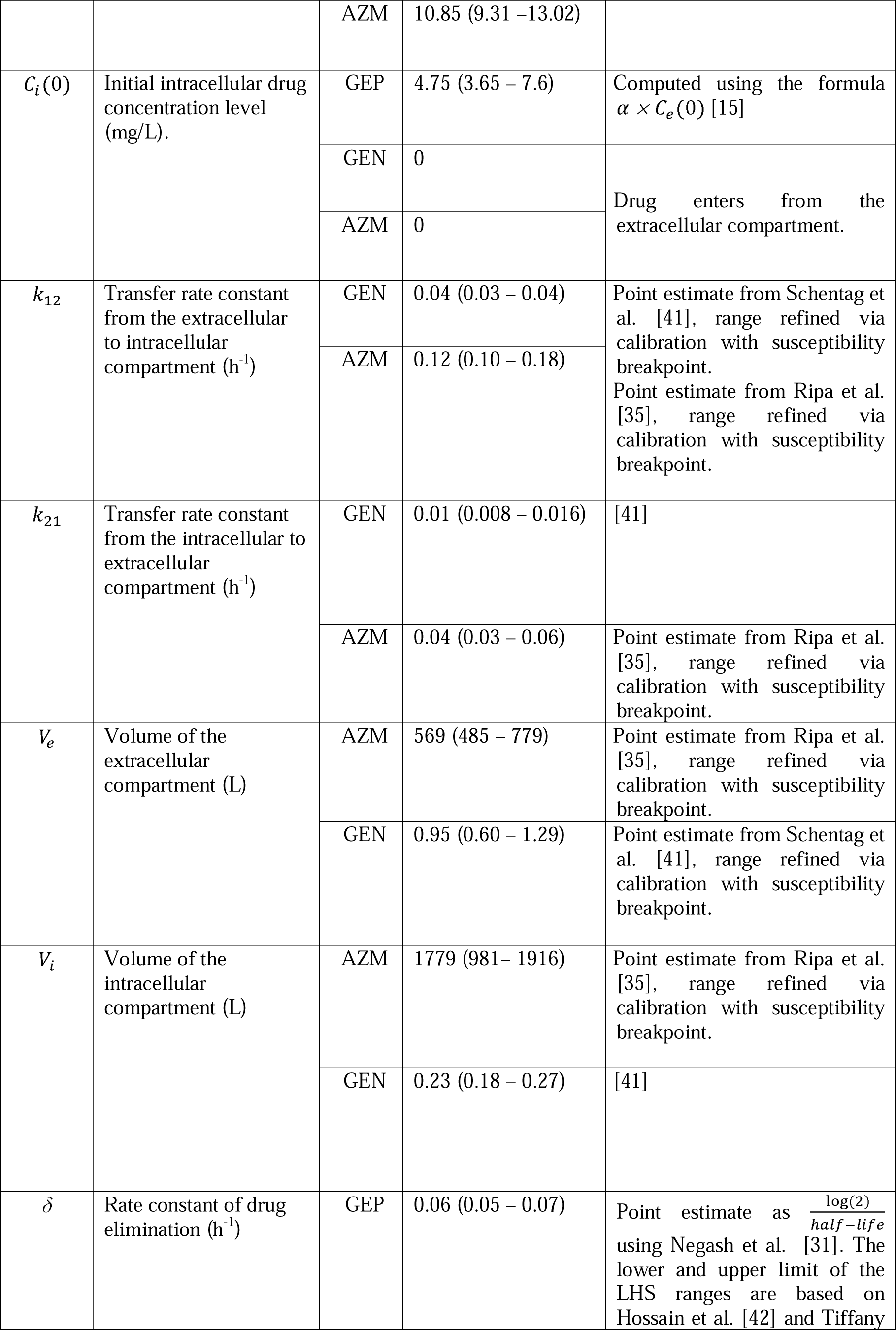
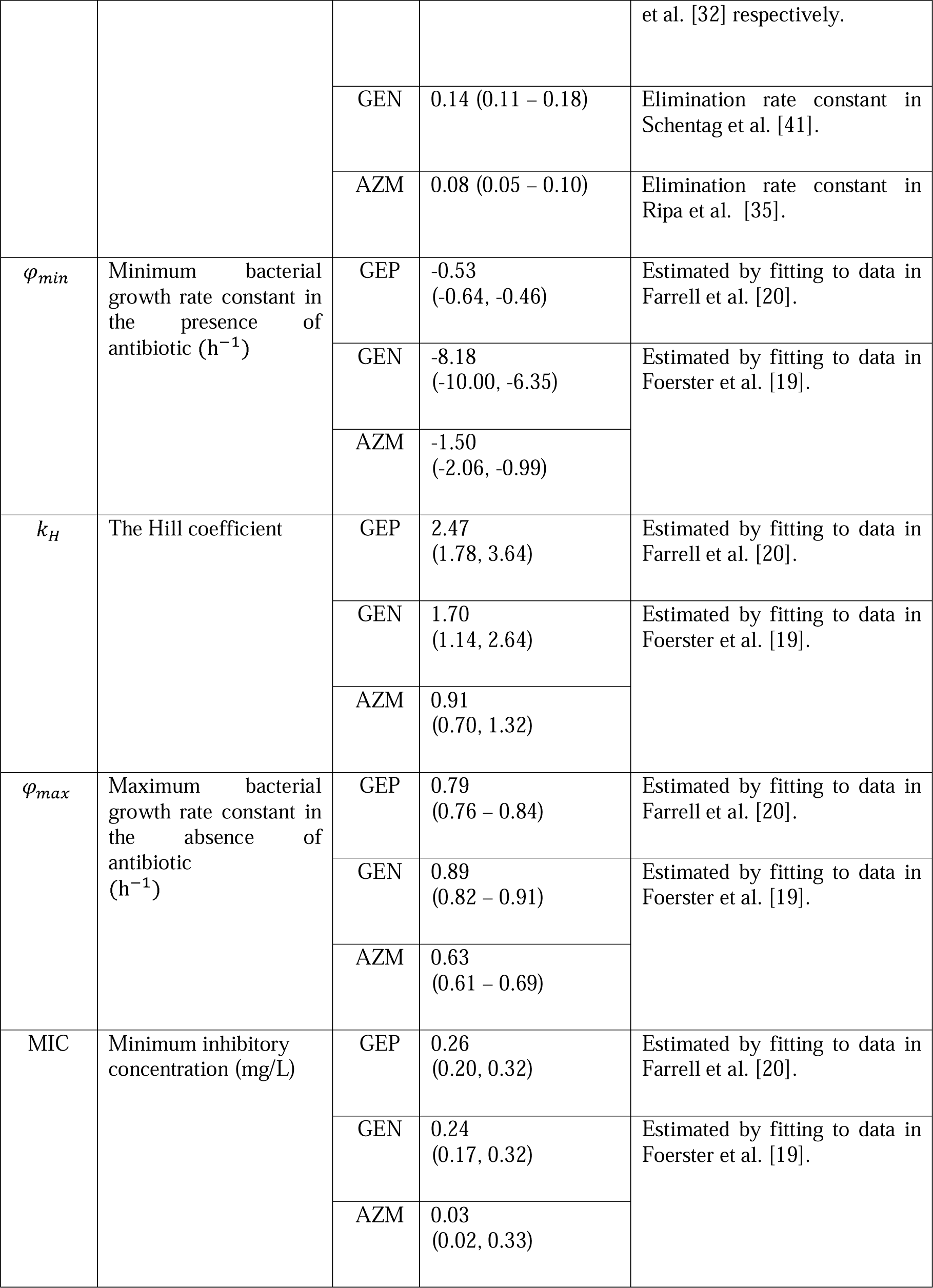
Model parameter values for the three antibiotics considered in this study: gepotidacin (GEP), gentamicin (GEN) and azithromycin (AZM).

| Symbol | Parameter (units) | Drug | Point Estimate (LHS range) | References/Comments |
| --- | --- | --- | --- | --- |
| $D$ | Initial antibiotic dose (mg) | GEP | 1500 / 3000 | Trial doses [5, 6]. |
|  |  | GEN | 240 | Trial doses [28, 29]. |
|  |  | AZM | 1000 | CDC recommended dose for dual treatment [30]. |
| $b_a$ | Bioavailability | GEP | 0.44 (0.38 – 0.5) | [31, 32] |
|  |  | GEN | 1 | Given intramuscularly [33]. |
|  |  | AZM | 0.37 | [27] |
| $V_d$ | Volume of distribution (L) | GEP | 188.7 | [31] |
|  |  | GEN | 16.8 (10 - 20) | [34] |
|  |  | AZM | 3219 (1593 - 5475) | [35] |
| $f_u$ | Fraction unbound | GEN | 0.85 – 1 | [36] |
|  |  | AZM | 0.88 | [37] |
|  |  | GEP | 0.76 | [38] |
| $\alpha$ | The ratio of intracellular to extracellular drug concentration | GEP | 1.8 (1.5 – 2.5) | [39] |
| $C_e(0)$ | Initial extracellular drug concentration level (mg/L) | GEP | 2.64 (2.43 – 3.04) | Computed using the formula $\frac{D \times b_a \times f_u}{V_d}$ [40] |
|  |  | GEN | 14.29 (10.21 – 23.76) |  |
|  |  | AZM | 10.85 (9.31 – 13.02) |  |
| $C_i(0)$ | Initial intracellular drug concentration level (mg/L). | GEP | 4.75 (3.65 – 7.6) | Computed using the formula $\alpha \times C_e(0)$ [15] |
|  |  | GEN | 0 | Drug enters from the extracellular compartment. |
|  |  | AZM | 0 |  |
| $k_{12}$ | Transfer rate constant from the extracellular to intracellular compartment ($\text{h}^{-1}$) | GEN | 0.04 (0.03 – 0.04) | Point estimate from Schentag et al. [41], range refined via calibration with susceptibility breakpoint.<br>Point estimate from Ripa et al. [35], range refined via calibration with susceptibility breakpoint. |
|  |  | AZM | 0.12 (0.10 – 0.18) |  |
| $k_{21}$ | Transfer rate constant from the intracellular to extracellular compartment ($\text{h}^{-1}$) | GEN | 0.01 (0.008 – 0.016) | [41] |
|  |  | AZM | 0.04 (0.03 – 0.06) | Point estimate from Ripa et al. [35], range refined via calibration with susceptibility breakpoint. |
| $V_e$ | Volume of the extracellular compartment (L) | AZM | 569 (485 – 779) | Point estimate from Ripa et al. [35], range refined via calibration with susceptibility breakpoint. |
|  |  | GEN | 0.95 (0.60 – 1.29) | Point estimate from Schentag et al. [41], range refined via calibration with susceptibility breakpoint. |
| $V_i$ | Volume of the intracellular compartment (L) | AZM | 1779 (981– 1916) | Point estimate from Ripa et al. [35], range refined via calibration with susceptibility breakpoint. |
|  |  | GEN | 0.23 (0.18 – 0.27) | [41] |
| $\delta$ | Rate constant of drug elimination ($\text{h}^{-1}$) | GEP | 0.06 (0.05 – 0.07) | Point estimate as $\frac{\log(2)}{\text{half-life}}$ using Negash et al. [31]. The lower and upper limit of the LHS ranges are based on Hossain et al. [42] and Tiffany et al. [32] respectively. |
|  |  | GEN | 0.14 (0.11 – 0.18) | Elimination rate constant in Schentag et al. [41]. |
|  |  | AZM | 0.08 (0.05 – 0.10) | Elimination rate constant in Ripa et al. [35]. |
| $\varphi_{min}$ | Minimum bacterial growth rate constant in the presence of antibiotic ($\text{h}^{-1}$) | GEP | -0.53<br>(-0.64, -0.46) | Estimated by fitting to data in Farrell et al. [20]. |
|  |  | GEN | -8.18<br>(-10.00, -6.35) | Estimated by fitting to data in Foerster et al. [19]. |
|  |  | AZM | -1.50<br>(-2.06, -0.99) |  |
| $k_H$ | The Hill coefficient | GEP | 2.47<br>(1.78, 3.64) | Estimated by fitting to data in Farrell et al. [20]. |
|  |  | GEN | 1.70<br>(1.14, 2.64) | Estimated by fitting to data in Foerster et al. [19]. |
|  |  | AZM | 0.91<br>(0.70, 1.32) |  |
| $\varphi_{max}$ | Maximum bacterial growth rate constant in the absence of antibiotic ($\text{h}^{-1}$) | GEP | 0.79<br>(0.76 – 0.84) | Estimated by fitting to data in Farrell et al. [20]. |
|  |  | GEN | 0.89<br>(0.82 – 0.91) | Estimated by fitting to data in Foerster et al. [19]. |
|  |  | AZM | 0.63<br>(0.61 – 0.69) |  |
| MIC | Minimum inhibitory concentration (mg/L) | GEP | 0.26<br>(0.20, 0.32) | Estimated by fitting to data in Farrell et al. [20]. |
|  |  | GEN | 0.24<br>(0.17, 0.32) | Estimated by fitting to data in Foerster et al. [19]. |
|  |  | AZM | 0.03<br>(0.02, 0.33) |  |

### Incorporation of parametric uncertainty

To account for parametric uncertainty across the natural infection model, in Jayasundara et al. [16] we selected 5402 parameter sets, generated using Latin hypercube sampling (LHS), which met the relevant outcome criteria for the natural time-course of infection (here we index these LHS parameter sets as i = 1, 2, . . ., 5402). To incorporate parameter uncertainty that is related to treatment, we extend this previous analysis by also simulating from the ranges that are associated with the treatment parameters. We achieve this by first generating 5402 uniform LHS samples (indexed as j = 1, 2, . . ., 5402) for the PK/PD parameters using the parameter ranges derived from relevant literature and summarised in Table 1 and Appendix S1, Table S2. Then to incorporate both natural infection and treatment-related parametric uncertainty, the LHS parameter sets that satisfy the indexing i = j are combined to result in 5402 sets of parameter values. Using these 5402 samples, we assess the modelled infection clearance times.

### Calibrating PK/PD parameters using susceptibility breakpoints

Explicitly capturing the development of antibiotic resistance would require considerable model extension with very limited data availability. Therefore, in this study, rather than directly modelling processes relating to antibiotic resistance, we vary the MIC as a proxy for changes in the susceptibility to a given treatment [43, 44]. To capture the notion of decreased susceptibility (or increased resistance) to treatment, we explore the effect of treatment via the MIC parameter in the Hill function (from here on referred to simply as the ‘MIC’), which we increase gradually from the antibiotic-specific MIC values estimated as described in Section ‘Mathematical model of antibiotic treatment’ for a susceptible NG strain. To this end, we determine a ‘model-derived susceptibility breakpoint’ such that for MIC below and above the breakpoint, the infection clears in ≤7 days and >7 days, respectively (infection clearance threshold, as described in Section ‘Simulated treatment strategies’). These model-derived breakpoints were then calibrated to reproduce the empirical breakpoints and thereby refine the ranges of the parameters that are influential in determining the model-derived susceptibility breakpoints (details of calibration are provided in the Appendix S1, Section S3). Here, we define ‘empirical breakpoints’ as the relevant susceptibility breakpoints for azithromycin published by the Clinical and Laboratory Standards Institute (CLSI) (1mg/L [45]) and the European Committee on Antimicrobial Susceptibility Testing (EUCAST) (0.5mg/L [46]). For gentamicin, a susceptibility breakpoint of 4mg/L is defined based on epidemiological and clinical observations in Malawi as reported in the study by Brown et al. [47]. Furthermore, Brown et al. [47] defines intermediate susceptibility for gentamicin for MIC 8-16mg/L and resistance for MIC ≥32mg/L [47]. For gepotidacin which is not currently used in clinical practice, we use the breakpoints determined in the clinical trials conducted by Taylor et al. [5] and Scangarella-Oman et al. [6].

### Simulated treatment strategies

In this study, we simulate the effectiveness of the single and multiple dose treatment strategies summarised in Tables 3 and 4. Here, we consider strategies that have been previously tested in clinical trials and compare the simulated treatment effectiveness with clinical trial results as well as using the model to simulate the effectiveness of several novel multiple-dose strategies. Therefore, as previously tested strategies, for gepotidacin we analyse the effectiveness of 1500mg and 300mg single dose strategies which are tested in the clinical trials Taylor et al. [5] and Scangarella-Oman et al. [6]. For the dual treatment combination we test 240mg GEN + 1g AZM strategy tested in the clinical trial Kirkcaldy et al. [8] and 240mg GEN + 2g AZM strategy tested in Rob et al. [9].

**Table 2:** Susceptibility breakpoints (mg/L) derived from the three sub-models and the full model and comparison with empirical breakpoints.

| Drug | Susceptibility breakpoints (mg/L) |  |  |  |  |
| --- | --- | --- | --- | --- | --- |
|  | Model A | Model B | Model C | Full model point estimate (LHS range) | Empirical breakpoints |
| GEP | 2.55 | 0.79 | 0.73 | 0.64 (0.48 – 1.1) | Not available |
| AZM | 9.35 | 0.89 | 0.70 | 0.69 (0.55 – 1.29) | 0.5 [46], 1 [45] |
| GEN | 12.75 | 1.94 | 1.74 | 1.60 (1.51 – 5.54) | 4 [47]. |

**Table 3:**
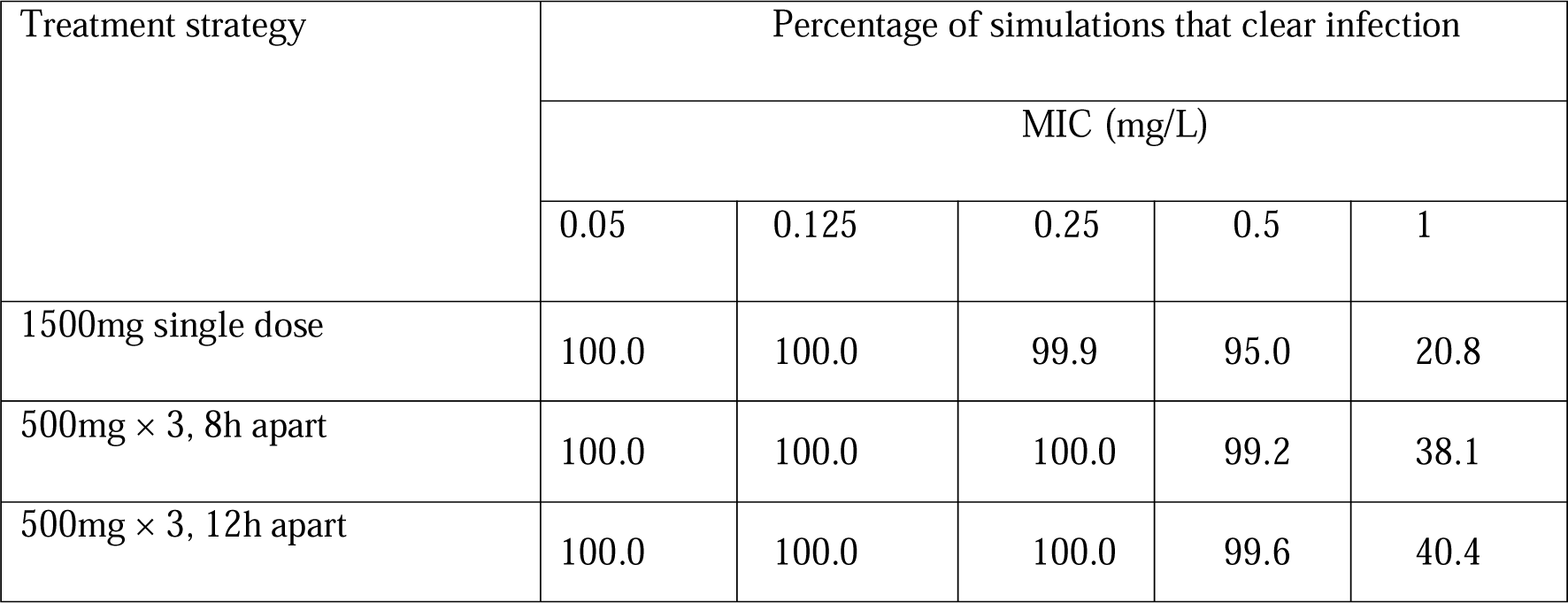
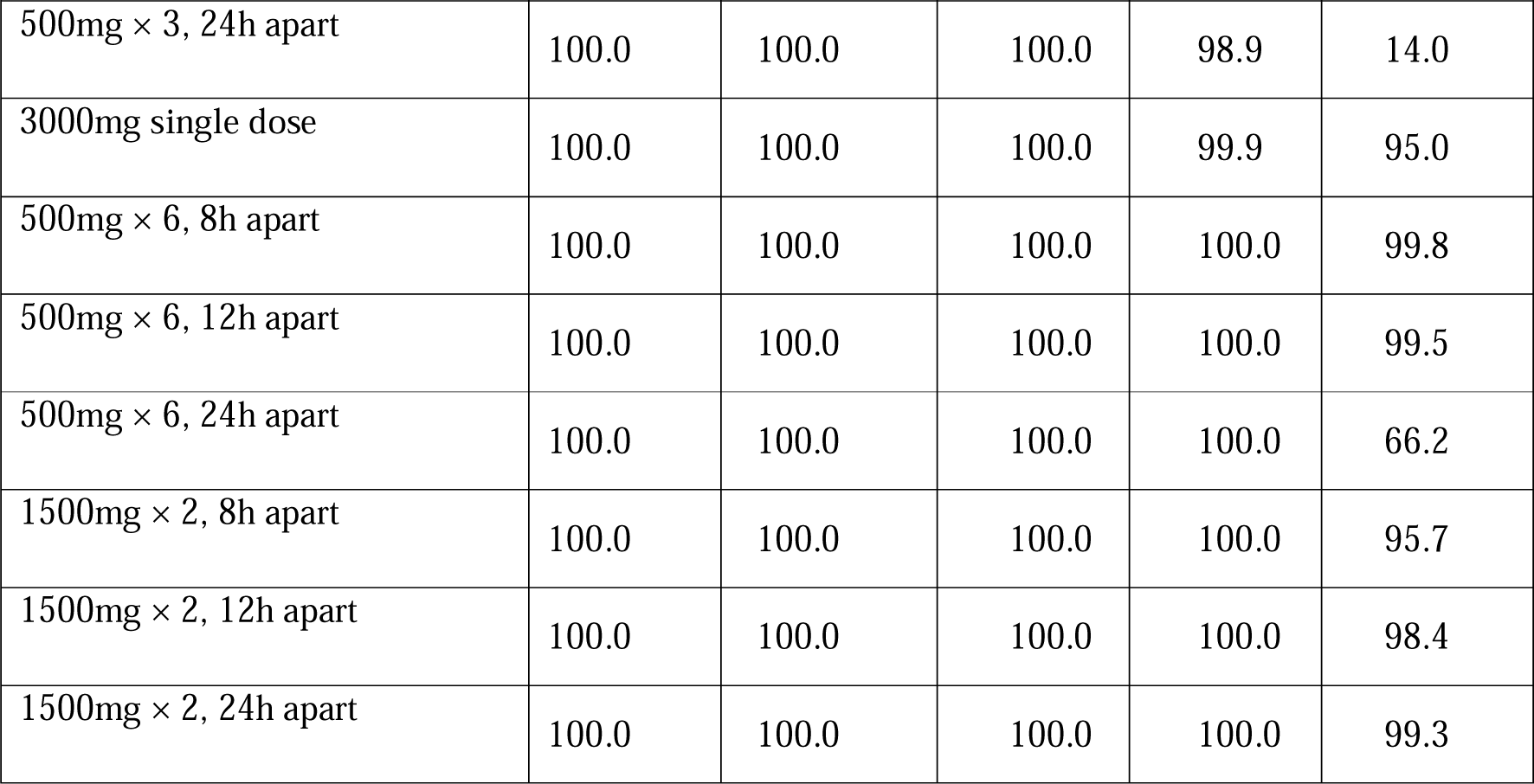
Percentage of simulations using LHS samples (out of 5402) that clear infection in ≤7 days when using single and multiple dose gepotidacin treatment strategies.

| Treatment strategy | Percentage of simulations that clear infection |  |  |  |  |
| --- | --- | --- | --- | --- | --- |
|  | MIC (mg/L) |  |  |  |  |
|  | 0.05 | 0.125 | 0.25 | 0.5 | 1 |
| 1500mg single dose | 100.0 | 100.0 | 99.9 | 95.0 | 20.8 |
| 500mg × 3, 8h apart | 100.0 | 100.0 | 100.0 | 99.2 | 38.1 |
| 500mg × 3, 12h apart | 100.0 | 100.0 | 100.0 | 99.6 | 40.4 |
| 500mg × 3, 24h apart | 100.0 | 100.0 | 100.0 | 98.9 | 14.0 |
| 3000mg single dose | 100.0 | 100.0 | 100.0 | 99.9 | 95.0 |
| 500mg × 6, 8h apart | 100.0 | 100.0 | 100.0 | 100.0 | 99.8 |
| 500mg × 6, 12h apart | 100.0 | 100.0 | 100.0 | 100.0 | 99.5 |
| 500mg × 6, 24h apart | 100.0 | 100.0 | 100.0 | 100.0 | 66.2 |
| 1500mg × 2, 8h apart | 100.0 | 100.0 | 100.0 | 100.0 | 95.7 |
| 1500mg × 2, 12h apart | 100.0 | 100.0 | 100.0 | 100.0 | 98.4 |
| 1500mg × 2, 24h apart | 100.0 | 100.0 | 100.0 | 100.0 | 99.3 |

**Table 4:** Percentage of simulations that clear the infection (out of 5402 LHS samples) at various MIC values with gentamicin (GEN) and azithromycin (AZM) dual therapy regimens.

| Treatment strategy | Percentage of simulations that clear infection |  |  |  |  |  |
| --- | --- | --- | --- | --- | --- | --- |
|  | (Gentamicin/azithromycin) MIC (mg/L) |  |  |  |  |  |
|  | (4/0.5) | (4/1) | (8/0.5) | (8/1) | (16/0.5) | (16/ 1) |
| **Strategies with gentamicin total accumulation of 240mg** |  |  |  |  |  |  |
| 240mg GEN + 1g AZM | 95.6 | 85.9 | 86.9 | 61.1 | 78.5 | 39.7 |
| 240mg GEN + 2g AZM | 99.7 | 95.6 | 98.8 | 86.9 | 97.7 | 78.5 |
| 80mg GEN × 3, 8h apart + 1g AZM single dose | 95.6 | 86.0 | 86.9 | 61.1 | 78.5 | 39.8 |
| 120mg GEN × 2, 8h apart + 1g AZM single dose | 95.6 | 86.2 | 86.9 | 61.9 | 78.8 | 39.8 |
| **Strategies with gentamicin total accumulation of 480mg** |  |  |  |  |  |  |
| 120mg GEN × 2, 12h apart for 2 days + 1g AZM single dose | 99.8 | 99.3 | 97.9 | 93.7 | 92.5 | 76.2 |
| 240mg GEN × 2, 24h apart + 1g AZM single dose | 99.8 | 99.2 | 97.8 | 93.1 | 92.1 | 74.8 |
| **Strategies with gentamicin total accumulation of 720mg** |  |  |  |  |  |  |
| 240mg GEN × 3, 24h apart + 1g AZM single dose | 99.9 | 99.7 | 98.8 | 96.4 | 94.5 | 82.2 |
| 240mg GEN × 3, 24h apart + 2g AZM single dose | 100.0 | 99.9 | 99.9 | 98.9 | 99.7 | 95.5 |
| 240mg GEN × 3, 24h apart + 1g AZM × 2, 24h apart | 100.0 | 99.9 | 99.8 | 98.5 | 99.4 | 93.5 |

Treatment is initiated at the peak NG load as identified in our model of untreated infection (at 3.6 days post-infection in the base case) [16], at which point we assume symptoms to be apparent. We classify simulations in which infection is cleared in ≤7 days as treatment success, as used in recent clinical trials [6, 48, 49] indicating this timeframe as appropriate to bound successful infection clearance. Simulated infections are assumed to be cleared when the total bacterial load (B+ B_a_ + B_i_ + B_S_) falls below 10 bacteria, as used in Jayasundara et al. [16].

Any regimen that is approved for the treatment of gonorrhoea should have ≥95% treatment efficacy [12, 50]. Here, we adopt an analogous definition in terms of our simulations whereby for a given MIC value if ≥95% of simulations that are generated from our LHS samples achieve treatment success we consider that particular treatment strategy to be effective. We henceforth define simulated ‘treatment effectiveness’ as the proportion of model simulations that result in successful infection clearance. We note that the sources of variation present in our model are not directly comparable to the variability observed during the treatment of natural human infection and these percentages cannot be directly interpreted as estimates of treatment effectiveness.

### Extracellular vs intracellular susceptibility breakpoints

To understand potential differences between *in vitro* and *in vivo* clearance behaviour, we compare the susceptibility breakpoints derived from sub-models of increasing complexity starting with only extracellular states and progressing to the full model involving epithelial cells and PMN.

Model A reflects an *in vitro* time-kill study, in which extracellular NG but no host cells (epithelial cells or PMN) are present. In simulations, NG are allowed to grow exponentially and the drug concentration is kept constant (no drug decay), similar to the experimental design used in the *in vitro* study by Foerster et al. [19]. In Model B, epithelial cells are added, leading to the inclusion of unattached NG, NG attached to epithelial cells and NG internalised within epithelial cells. In model C, NG interaction with epithelial cells is removed but the PMN response and NG survival within PMN are included in the simulations. In models B and C and the full-treatment model, logistic constraints on growth are applied as described previously in Jayasundara et al. [16] and the drug concentration varies over time as described above in Section ‘Mathematical model of antibiotic treatment’. Comparisons of the derived susceptibility breakpoints are then made between the sub-models and the full model for the same initial extracellular drug concentration.

### PK indices

To compare the effectiveness of differing gepotidacin treatment regimens, we evaluate three PK indices: time above the MIC (t_MIC_); the ratio of area under the drug concentration curve to the MIC (AUC/MIC); and the ratio of peak drug concentration to the MIC (C_max_/MIC). The area integrated over the total drug concentration curve (AUC_O-oo_/MIC) is used as the default AUC/MIC index but we also test the area under the curve above the MIC (removing the area below the MIC from the total area under the curve) and AUC over a fixed time period of 7 days (AUC_O-7_/MIC) as alternative indices (see Appendix S1, Section S7.3). For multiple dose strategies, we also calculate the total time the drug concentration remains above the MIC (t_MIC_) and this is used as the default index of t_MIC_, and additionally consider some alternative definitions of t_MIC_ in the Appendix S1, Section S7.3. We calculate the three PK indices separately for intracellular and extracellular drug concentrations labelling these indices with the subscripts ‘in’ and ‘ex’ (e.g., t_MIC_in__, t_MIC_ex__).

Similarly, for the dual treatment option we calculate the ratio of area under the drug concentration curve to the MIC (AUC/MIC_h_) using the simulated single drug concentration representing the combined effect of gentamicin and azithromycin calculated using the Loewe additivity concept (using Appendix S1, Equation S4). Loewe additivity combines both antibiotics, gentamicin and azithromycin into a single drug of higher effect. Here, MIC_h_ refers to the MIC of the drug having the higher effectiveness out of gentamicin (4mg/L) and azithromycin (1mg/L) at each time point. This PK index is calculated in both the extracellular and intracellular environments and a threshold is determined to distinguish treatment success and failure.

### Non-adherence to treatment strategies

For multiple dose strategies of gentamicin which extend over 3 days, we also test the impact of limited non-adherence by the patient. Specifically, we consider a uniformly distributed delay of between 0 and 24h to the 2^nd^ dose in comparison to the recommended schedule, with subsequent doses then taken at the correct spacing from the previous dose. Treatment efficacy is analysed when 15%, 25% 50%, 75% and 100% of the simulations deriving from the LHS samples are assumed to be subject to non-adherence.

## Results

### Extracellular vs intracellular susceptibility breakpoint

For each of the sub-models described in Section ‘Simulated treatment strategies’ we determine drug-specific model-derived susceptibility breakpoints, with simulation results based on point estimates summarised together with those from the full treatment model in Table 2. In addition, breakpoint ranges derived from simulations using all LHS parameters are provided for the full model and compared with empirical breakpoints where available.

We observe that with the addition of intracellular compartments the model-derived susceptibility breakpoints are 8-fold, 14-fold and 4-fold lower in the full model as compared to the *in-vitro* model (model A) for azithromycin, gentamicin and gepotidacin respectively. Results for models B (unattached and attached NG and NG within epithelial cells) and C (unattached NG and NG within PMN) are similar to those for the full model, indicating that these large differences in model-derived susceptibility breakpoints for model A compared with the other models is associated with the inclusion of intracellular NG states in simulations.

### Gepotidacin monotreatment

The results of model simulations for gepotidacin monotreatment are summarised in Table 3. Gepotidacin regimens that accumulate to 1500mg in total, irrespective of administration as single or multiple doses, achieve treatment success for NG MIC ≤0.5mg/L, while most regimens with a total dose of 3000mg achieve success for MIC ≤1mg/L. In our model, clearance behaviour is invariant when the MIC/dose ratio is held fixed (see Appendix S1, Section S7.1), with higher dose strategies of 4.5g and 6g gepotidacin being successful for MIC ≤1.5mg/L and MIC ≤2mg/L, respectively (Appendix S1, Table S3).

Some of the multiple dose regimens for gepotidacin we investigate have not yet been tested in clinical trials. In the majority of simulated regimens, treatment success/failure is consistent across single and multiple dose strategies with the same total dose amount for the same NG MIC parameter. However, daily administration of 500mg for 6 days at MIC=1mg/L, resulted in treatment failure (∼66% of simulations cleared), despite treatment success with other 3000mg total dose regimens simulated here. We discuss this result in more detail in the next section.

### Effectiveness of different dosing strategies of gepotidacin

Comparison of PK indices across the gepotidacin regimens provides insight into why the simulated 500mg × 6, at 24h interval regimen failed treatment at MIC=1mg/L whereas other regimens with the same total drug did not. In this regimen, the intracellular drug concentration was maintained above the MIC (t_MIC_) for only 47% of the dosing interval and correspondingly bacterial load spiked as the drug concentration fell below the MIC (Fig. 2). By comparison, for 500mg × 6 dosing regimens at intervals of 8 and 12h the intracellular drug concentration is above 1mg/L for 100% and 94% of the dosing interval, respectively.

**Fig 2:**
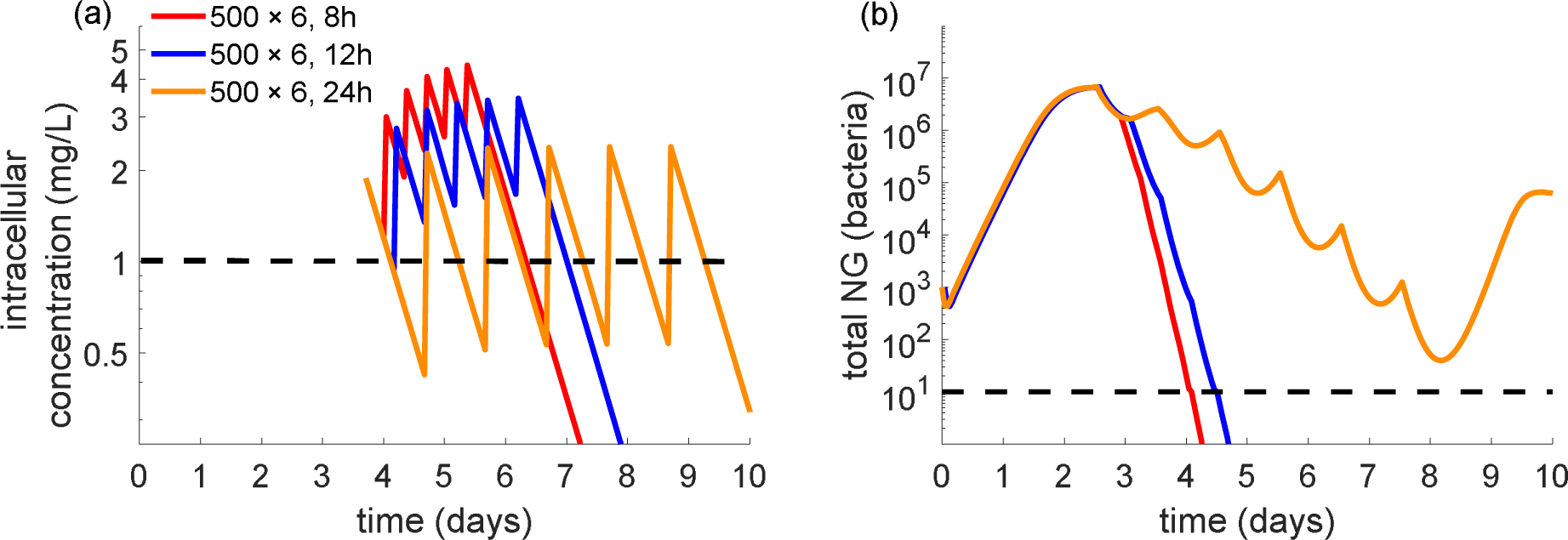
Effect of gepotidacin dosing intervals of 8,12 and 24h in a 500mg × 6 schedule on (a) intracellular drug concentration and (b) total NG load. Dashed lines indicate MIC of 1mg/L (a) and infection clearance cut-off of 10 bacteria (b). Parameter values are specified in Table 1.

The simulation results in Table 3 also suggest that in most cases, multiple dose regimens clear infection in a higher fraction of simulations when the total dose is held fixed. For instance, at a MIC for gepotidacin of 0.5mg/L infection clearance occurs in 95.0% of simulations with a 1500mg single dose compared with >98% simulations in 500mg × 3 regimens at 8, 12 and 24h intervals. Here, the multiple dose strategies achieve an increased t_MIC_ in comparison to the single dose strategy (Appendix S1, Fig. S7). The highest value of this PK index also occurs with the most effective dosing interval (24h) at MIC of 1mg/L with a total dose of 3000mg split into two (1500mg × 2 given 8, 12 or 24h apart) as shown in Appendix S1, Fig. S7.

### PK indices to differentiate treatment success using gepotidacin

We also attempt to determine treatment success and failure based on PK indices evaluated using extracellular and intracellular gepotidacin concentration. Extracellular PK indices fail to sharply distinguish simulations in which treatment succeeds from those where it fails, as there are simulations with the same PK index value but opposite treatment outcomes (Fig.3). The ratio of peak intracellular drug concentration to MIC (C_max_/MIC_in_) index is also unable to discriminate between success or failure to clear infection. In contrast, intracellular indices for the ratio of area under the total drug concentration curve to the MIC (AUC/MIC_in_) and time above the MIC (t_MIC_), clearly differentiate between treatment success and failure. However, while a common cut-off across all dosing schedules could be obtained with the AUC/MIC_in_ index (Fig.3), the t_MIC_ cut-off varies by dosing schedule. This behaviour is preserved under the alternative definition whereby only the AUC above the MIC is considered (Appendix S1, Fig. S9). Dose-dependence also occurs for other forms of the t_MIC_ cut-off (Appendix S1, Fig. S8). We therefore focus on the AUC/MIC_in_ index for gepotidacin in regard to determination of a threshold parameter.

**Fig 3:**
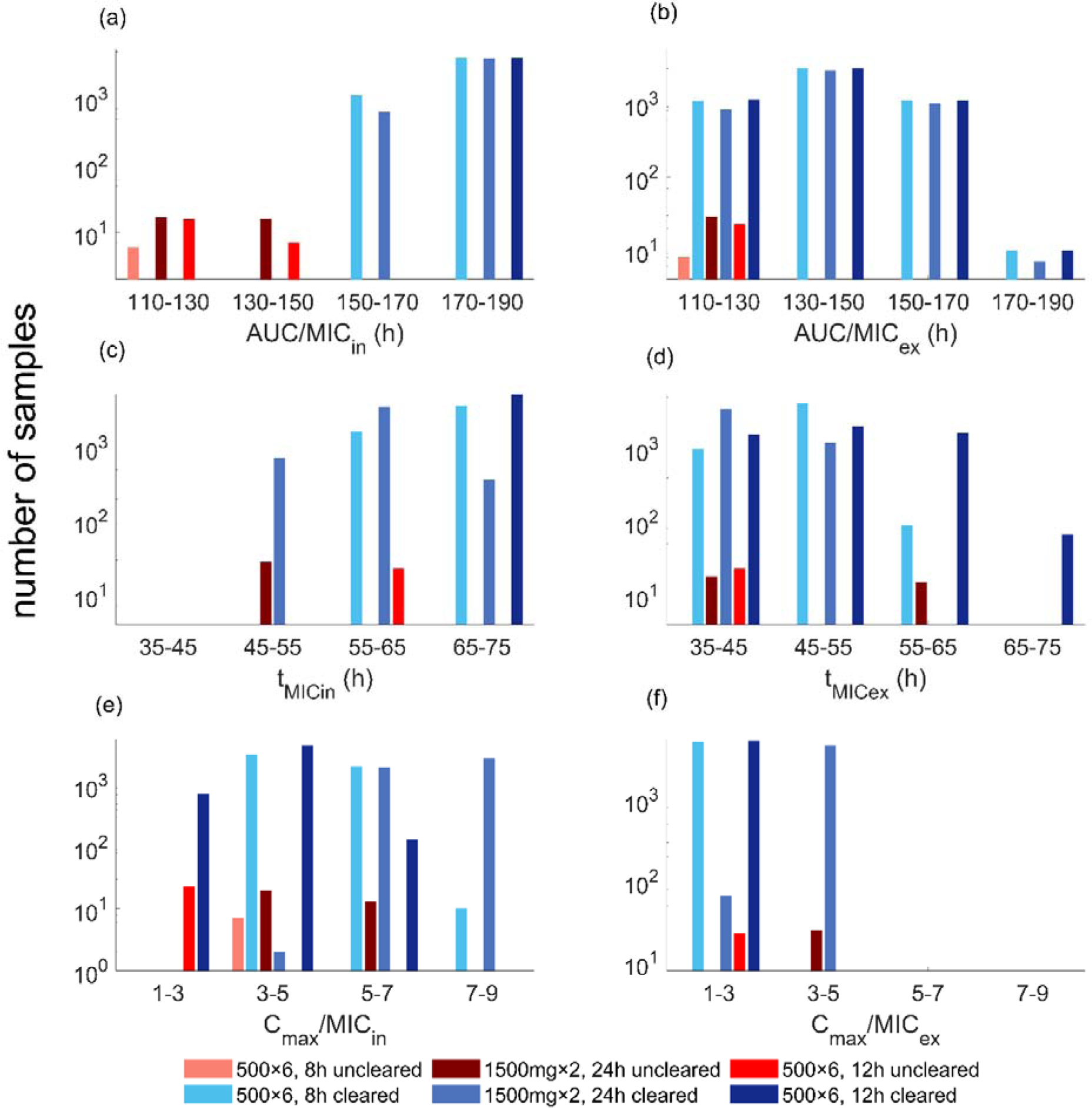
Comparison of PK/PD indices to differentiate treatment success and failure. The ratio of area under the curve to the MIC are shown for: (a) intracellular and (b) extracellular drug concentration; the time above the MIC calculated for intracellular (c) and extracellular (d) drug concentration; the ratio of peak drug concentration to the MIC for intracellular (e) and extracellular (f) drug concentration.

From the simulated concentration profiles, we observe that treatment success for gepotidacin occurs in simulations where AUC/MIC_in_ >150h (Fig.3). We note that there are 6 simulations with AUC/MIC_in_ in the range of 147-150h that fail to clear the infection (simulation behaviour shown in Appendix S1, Fig. S10). For these unsuccessful simulations, the total bacterial load declines very close to the infection clearance threshold (to ∼11 bacteria in some instances), but does not meet our criterion for infection clearance (total NG load <10 bacteria). This further supports the AUC/MIC_in_ >150h, as a suitable threshold to differentiate between simulated treatment success and failure.

### Dual treatment with gentamicin + azithromycin

#### Effectiveness of different dosing strategies of gentamicin + azithromycin

The effectiveness of dual treatment with gentamicin + azithromycin across single and multiple dose strategies is summarised in Table 4. For the same total dose amount, multiple doses of gentamicin and multiple doses of azithromycin result in similar effectiveness to the single dose strategy, with limited sensitivity to dosing frequency as well. Among the tested strategies, only 240mg × 3 gentamicin, given 24h apart in combination with 2g single dose of azithromycin is effective at high MIC for both gentamicin and azithromycin (16 mg/L and 1 mg/L, respectively, Table 4). We also examine the impact of limited non-adherence using the multiple dose strategy of 240mg × 3 gentamicin, given 24h apart along with 2g single dose of azithromycin. Here, at MIC for gentamicin and azithromycin of 16mg/L and 1 mg/L, respectively, for the 100% non-adherence scenario 94.13% (Appendix S1, Table S4) treatment success is observed showing similar effectiveness (95.45%) to the 100% adherent scenario (Table 4).

#### PK index to differentiate treatment success using the dual treatment combination of gentamicin and azithromycin

We also attempt to distinguish treatment success and failure based on the PK index evaluated using the single drug resulting from Loewe additivity. Similar to gepotidacin, the ratio of area under the total intracellular drug concentration curve to the MIC (AUC/MIC_h,in_) can clearly differentiate between treatment success and failure (Fig. 4). From the simulated concentration profile of a single drug resulting from Loewe additivity, we observe that all samples achieving AUC/MIC_h,in_ > 140h successfully clear infection. However, unlike the PK index threshold related to gepotidacin monotreatment, we observe that a substantial proportion of simulations successfully clear infection when AUC/MIC_h,in_ < 140h.

**Fig 4:**
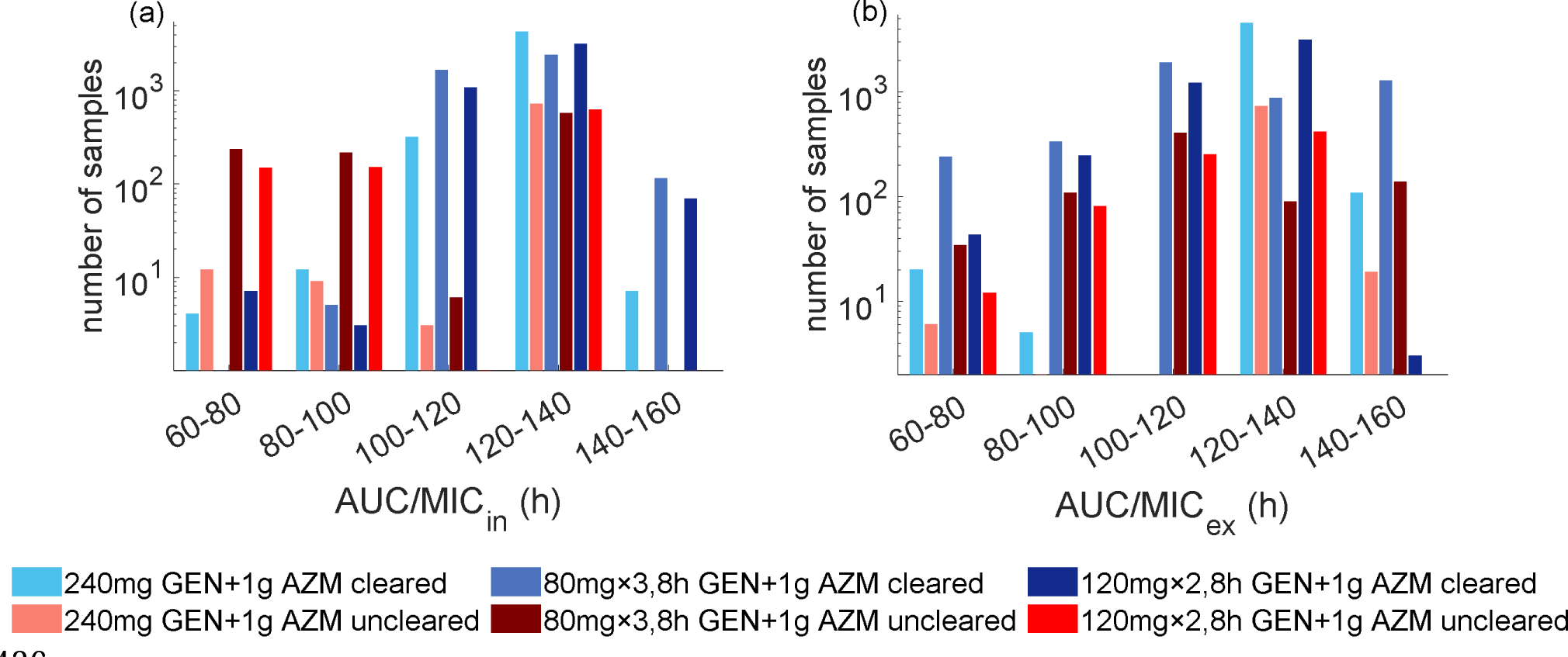
Simulated infection clearance based on the ratio of area under the (a) intracellular and (b) extracellular drug concentration resulting from Loewe additivity (AUC/MICh) for gentamicin and azithromycin dual treatment option.

## Discussion

In this study, we develop a within-host mathematical model to describe antibiotic treatment effects while considering NG interactions with host cells. We found that inclusion of intracellular states leads to substantial changes in MIC clearance thresholds as opposed to *in-vitro* NG dynamics alone. The relevance of different intracellular NG states in determining treatment success is a matter of current debate by experts in this field [51]. The difficulty in reaching a consensus on this issue is likely due to limited experimental evidence of the impact of intracellular antibiotic-mediated killing on treatment outcomes. Here, our findings on the model-derived susceptibility breakpoints and treatment effects in the presence of intracellular NG, suggest further experiments assessing the role of intracellular NG in determining treatment success could be valuable. We also analyse the association of PK indices with treatment success and the level of intracellular drug concentration that must be maintained to achieve successful infection clearance. When calculating PK indices relevant to the dual treatment option we introduce a novel approach of using a simulated single drug concentration representing the combined effect of gentamicin and azithromycin calculated using the Loewe additivity concept. However, unlike in the monotreatment case, the threshold relating to dual treatment does not separate treatment success from treatment failure as a majority of samples below our PK index threshold still lead to clearance.

In Jayasundara et al. [16], we showed the importance of intracellular NG in prolonging the duration of natural infection and here we show the importance of intracellular antibiotic mediated killing in determining treatment success in our model. The importance of different intracellular NG states (NG within PMN and epithelial cells) in determining treatment success is not as yet resolved [51], due to limited experimental evidence of the impact of intracellular antibiotic-mediated killing on treatment outcomes. Although *in vitro* models such as those developed using immortal cell lines (e.g., HeLa cells) [52] have been used to explore the intracellular behaviour of NG, we are not aware of any study that considers antibiotic interactions with intracellular NG. Here, our findings on the model-derived susceptibility breakpoints in the presence of intracellular NG, suggest further experiments assessing the role of intracellular NG in determining treatment success could be valuable.

Building on our simulation results highlighting the importance of intracellular concentrations in treatment success, we found that an intracellular version of the area under the curve index discriminated between treatment success and failure using gepotidacin. Consistent with our findings, a strong correlation between AUC/MIC index and bacterial killing of two gram-positive pathogens (*S.aures* and *S.pneumoniae*) has been reported by Bulik et al. [38]. Although our extracellular index measures align with the calculations based on plasma drug concentrations in Scangarella-Oman et al. [6], their study is limited by sample size with only five NG isolates with MIC for gepotidacin of 1mg/L [53]. However, in our model treatment success and failure could only be clearly differentiated through intracellular indices. This is because, in our model implementation, consistent with limited empirical evidence of intracellular NG populations measured in urethral exudates by Veale et al. [54], a majority of NG reside intracellularly [16] and here, treatment success is observed to be mainly determined through the killing of intracellular NG (Table 2).

Our analysis of dual treatment using single doses of gentamicin + azithromycin is comparable, to a certain extent, with the limited data available from clinical trials. The two clinical trials that have been conducted for this drug combination report an overall genital infection treatment success rate of 94% [7] and 100% [8] using 240mg gentamicin combined with 1g and 2g azithromycin doses, respectively. In the clinical trial by Ross et al. [7], 97.7% and 95.7% of isolates had MIC for gentamicin ≤4mg/L and MIC for azithromycin ≤0.5mg/L, respectively. However, in these studies treatment success is not disaggregated into MIC ranges and therefore, a clear comparison cannot be made with our model simulation results for MIC for gentamicin and azithromycin of 4mg/L and 0.5mg/L, respectively.

If additional data on antibiotic mediated killing of intracellular NG become available through future experimental studies, analogous for example to the *in vitro* time-kill experiment by Barcia-Macay et al. [55] that analysed drug mediated killing of extracellular and intracellular *S. aureus,* some of our findings on the rates of intracellular NG killing by antibiotics could then be compared with experimental data. Although such experimental studies on antibiotic activity against other intracellular pathogens can be a useful guide it is important to note that the magnitude of intracellular bacteriostatic/bactericidal effects depends on both the pathogen and the drug [56].

While most PK parameters (e.g., volume of distribution, drug half-life) are based on plasma drug concentration profiles measured in patients, we have had to rely on *in vitro* data for the PD parameters and some PK parameters. The experimental limitations of these *in vitro* studies, such as the use of constant drug concentrations and lack of intracellular bacteria, do not reflect the true *in vivo* environment and add potential for error in these parameters. Reflecting the limited data available, we took a parsimonious approach in assuming that intracellular PK effects for PMN and epithelial cells were the same. Although we recognise that both drug accumulation and penetration can depend on the host cell and tissue type [56, 57] we lacked relevant data to inform different estimates.

## Conclusions

In this study, we developed a PK/PD analysis approach to study antibiotic interaction with NG in different cellular states and to assess the effectiveness of novel treatment strategies over a range of MIC values. To the best of our knowledge, this is the first within-host mathematical modelling study that explores the intracellular antibiotic killing of NG. Our findings suggest the importance of considering intracellular dynamics when deciding on treatment regimens as the model-derived susceptibility breakpoints are observed to be substantially impacted by the killing of NG within PMN and epithelial cells. This also draws attention to the potential importance of further experimental studies that capture intracellular PK/PD effects in regard to gonorrhoea treatment. Such investigation into the intracellular antibiotic effects may be useful when developing novel antibiotics for gonorrhoea. In addition, our findings, and the model more generally, may have utility as a tool for identifying treatment regimens to explore further in clinical trials.

## Data Availability

All relevant data are within the paper and its Supporting Information files and the code is available on GitHub at https://github.com/pavijayasundara/NG-Treatment-Model.

## Author contributions

P.J conceptualisation, study design, model development, validation, figures, and writing. JGW, DGR, PK methodology, study design, review, editing, supervision. All authors reviewed the manuscript.

## Competing Interests

Author’s declare no competing interests. This work was supported by funding from the National Health and Medical Research Council (NHMRC) [grant numbers APP1078068 and APP1071269]. PJ was supported by a UNSW Sydney PhD tuition fee scholarship.

## Notes

### Competing Interest Statement

The authors have declared no competing interest.

### Author Declarations

The study used only openly available data and data sources are provided in the manuscript.

