## Supplementary File for "Modelling treatment effects for gonorrhoea"

### Appendix S1

#### S1 Modelling pharmacodynamics.

##### S1.1 The Hill function

Drug effects on the NG population are modelled using a Hill function as in Regoes et al., [1], where the authors used the Hill function to describe the relationship between the growth rates of *E.coli* and the concentrations of antibiotics (*C*) of different classes that are measured through *in vitro* time-kill experiments. The Hill function is determined by four parameters: the maximum ($\varphi_{max}$) and minimum ($\varphi_{min}$) bacterial growth rates in the absence and presence of the antibiotic, respectively; the MIC; and the Hill coefficient ($k_{H}$), which reflects the sensitivity of the change in the net bacterial growth rate to the changes in the antibiotic concentration $\varphi\left( C \right)$. Under this parameterisation, $\varphi\left( C \right)$ is then described by Eq. S1:

$$\varphi\left( C \right)=\varphi_{max}-\frac{\left( \varphi_{max}-\varphi_{min} \right)\left( \frac{C}{MIC} \right)^{k_{H}}}{\left( \frac{C}{MIC} \right)^{k_{H}} - \frac{\varphi_{min}}{\varphi_{max}}}. (S1)$$

##### S1.2 Data used to estimate Hill function parameter values

The Hill function parameters are estimated using the NG growth data reported in the *in vitro* time-kill experiments for gentamicin and azithromycin in the study by Foerster et al., [2] and for gepotidacin in the study by Farrell et al., [3]. These studies measure bacterial growth at various time points in the absence and presence of the antibiotic at a stable concentration using wild-type NG strains that do not express resistance against the tested antibiotics. Foerster et al., [2] reports drug concentrations in the range of 0.016 × MIC to 16× MIC while the study Farrell et al., [3] reports drug concentrations ranging from 0.25 × MIC to 10 × MIC. Foerster et al., [2] have conducted two identical experiments on NG growth and we consider both these experiments when estimating the parameter values. In Foerster et al., [2] NG count below 100 CFU/mL could not be measured, so only the concentrations with at least three data points are used for the optimisation. For gentamicin, the number of antibiotic concentrations that are used in the optimisation (*n*) is 8 in each experiment while for azithromycin *n* is 11 in each. In Foerster et al., [2], NG growth is measured hourly for 6 hours (0, 1, 2, 3, 4, 5, 6 hour time points) and in the study Farrell et al., [3] it is measured at 0, 2, 4, 8 and 24 hour time points. As in Foerster et al., [2], the geometric mean of NG data at 0 hours is used as the initial bacterial load (at 0 hours).

##### S1.3 Hill function parameter estimation.

In Foerster et al., [2] NG growth data are presented for two separate experiments. Therefore, in order to obtain the point estimate model parameter values an additional step is taken. First, the Hill function parameters are estimated by fitting separately to the two experimental data sets. This results in two sets of Hill function parameter values. Using these estimated individual sets of parameter values the net growth rates are evaluated using which we then evaluate the mean net growth rate. The Hill function is then fitted to this obtained mean net growth to estimate point estimate parameter values. These individual net growth rates and the fit to mean net growth rate are shown in Figure S1 for all the nine antibiotics studied in Foerster et al., [2]. The estimated individual parameter values are summarised in Table S1 along with the sum of squared errors obtained using our approach and using the estimates in Foerster et al., [2].

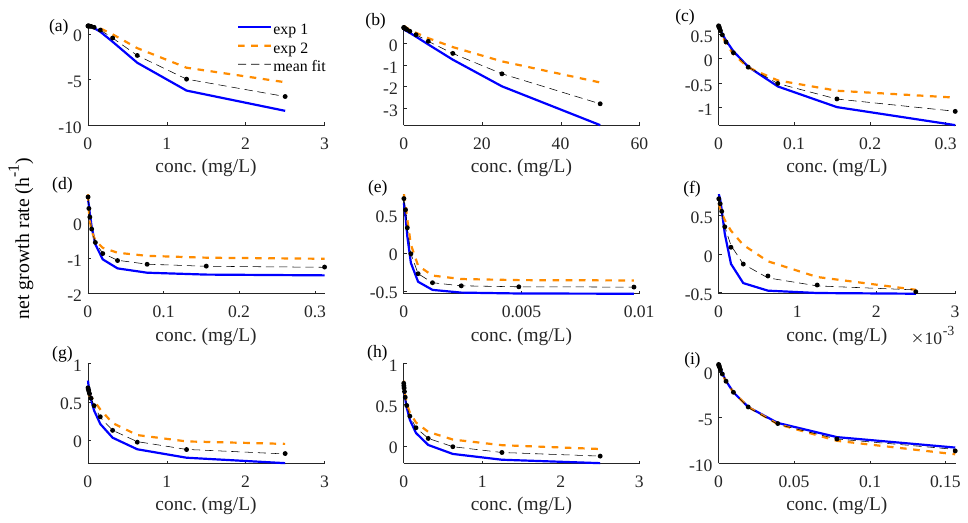

Figure S1: The net growth rates obtained using the individual parameter values obtained by fitting to the two experimental data sets in Foerster et al., [2] and the fit to the mean of the individual net growth rates (filled circles indicate data points for mean net growth rate and the dashed line represents the fit) for (a) gentamicin, (b) spectinomycin, (c) azithromycin, (d) penicillin, (e) ceftriaxone, (f) cefixime, (g) chloramphenicol, (h) tetracycline and (i) ciprofloxacin.

Table S1: The Hill function parameter values obtained by fitting to individual experiments and comparison of SSE using our estimates and Foerster et al., [2] estimates.

| Drug^a^ | Experiment | $\varphi_{max}$  (*h*^-1^) | $\varphi_{min}$ (*h*^-1^) | MIC (mg/L) | $k_{H}$ | Sum of squared errors (SSE) | SSE using estimates in Foerster, Unemo (2) |
| --- | --- | --- | --- | --- | --- | --- | --- |
| GEN | 1 | 0.82 | -10 | 0.19 | 1.67 | 3.11 | 5.82 |
|  | 2 | 0.91 | -6.35 | 0.3 | 1.76 | 4.86 | 10.59 |
| SPT | 1 | 0.66 | -10 | 5.96 | 1.12 | 5.76 | 9.66 |
|  | 2 | 0.85 | -10 | 10 | 0.83 | 3.99 | 13.95 |
| AZM | 1 | 0.61 | -0.99 | 0.02 | 0.97 | 10.47 | 45.6 |
|  | 2 | 0.69 | -2.06 | 0.03 | 0.91 | 5.29 | 26.02 |
| PEN | 1 | 0.67 | -1.48 | 4.20 × 10^-3^ | 1.35 | 11.27 | 16.16 |
|  | 2 | 0.92 | -1.05 | 2.80 × 10^-3^ | 0.86 | 19.05 | 21.39 |
| CFO | 1 | 0.68 | -0.52 | 2.42 × 10^-4^ | 1.74 | 6.08 | 9.40 |
|  | 2 | 0.77 | -0.36 | 3.84 × 10^-4^ | 1.76 | 6.94 | 7.50 |
| CFM | 1 | 0.78 | -0.52 | 1.23 × 10^-4^ | 1.82 | 9.95 | 12.30 |
|  | 2 | 0.67 | -0.76 | 4.63 × 10^-4^ | 0.87 | 8.77 | 13.81 |
| CHL | 1 | 0.77 | -0.04 | 0.35 | 0.89 | 5.32 | 7.58 |
|  | 2 | 0.59 | -0.07 | 1.00 | 1.64 | 2.04 | 2.36 |
| TET | 1 | 0.73 | -0.26 | 0.33 | 0.94 | 2.69 | 2.84 |
|  | 2 | 0.80 | -0.12 | 1.37 | 0.77 | 2.01 | 2.34 |
| CIP | 1 | 0.90 | -6.62 | 1.9 × 10^-3^ | 0.95 | 2.45 | 5.30 |
|  | 2 | 0.73 | -5.5 | 2.0 × 10^-3^ | 1.21 | 17.12 | 22.42 |

^a^GEN – gentamicin, SPT – spectinomycin, AZM – azithromycin, PEN – penicillin, CFO – ceftriaxone, CFM – cefixime, CHL – chloramphenicol, TET – tetracycline, CIP – ciprofloxacin.

###### S1.3.1 Parameter ranges around the Hill function estimates azithromycin and gentamicin.

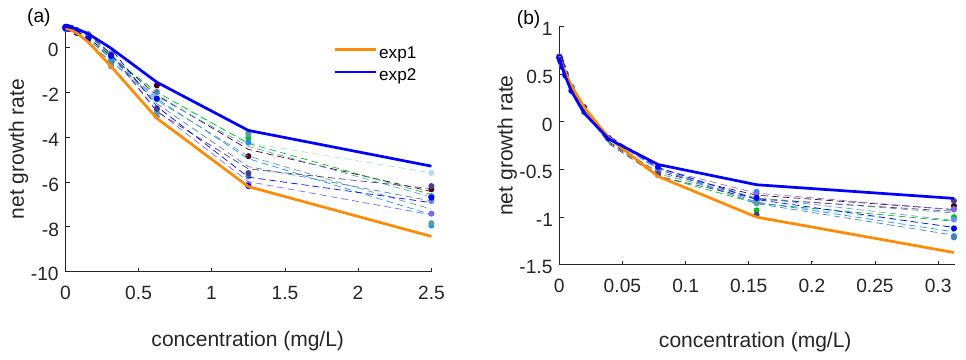
 Assuming that the resulting net growth rates can show any behaviour between the two individual net growth rates, in order to obtain parameter ranges around the point estimate Hill function parameters, a similar approach as fitting to the mean of the net growth rates as described in Section S1.3 is used. These parameter ranges are used in the uncertainty analysis described in Section 2.2 of main text. To estimate parameter ranges, for a particular antibiotic, at each concentration, 5402 uniform random numbers are generated between the two individual net growth rates (~Uniform ($\varphi_{1}(C),\varphi_{2}(C)$)), where $\varphi_{i}(C)$ are the respective individual net growth rates of the two experiments at concentration $C$, where *i=*1, 2. To each of these generated net growth rate curves, the Hill function (Eq. S1) is fitted to obtain 5402 sets of parameter values of $\varphi_{max}$, $\varphi_{min}$, MIC and $k_{H}$ for ceftriaxone, cefixime, azithromycin and gentamicin (Fig. S2).

Figure S2: Fits to generated net growth rates to estimate parameter ranges around the point estimate Hill function parameter values for (a) gentamicin and (b) azithromycin. Only 10 samples of the generated net growth rates (filled circles) and the fit of the Hill function to these data points (dotted lines) are shown. The solid lines indicate the individual net growth rates obtained using the two experimental NG load data in Foerster et al., [2].

#### S2 Modelling pharmacokinetics

##### S2.1 Modelling gepotidacin pharmacokinetics

When modelling gepotidacin concentration, we adopt a one-compartment model [4] as has been applied by So et al., [5] where we assume that drug concentration declines exponentially on a time-scale determined by the half-life of the drug. Specifically, extracellular drug concentration is modelled as $\frac{dC_{e}}{dt}\text{ = - }\text{δ}C_{e},$where $\text{δ}$ is the rate of decline of drug concentration and $C_{e}$ is the extracellular drug concentration. Here, only the free drug (drug unbound to albumin) is considered in the analysis as only the free drug molecules are considered to be microbiologically active [6, 7].

As gepotidacin show relatively low intracellular accumulation compared with drugs such as azithromycin [8, 9], we assume the intracellular concentration to be proportional to the extracellular concentration with proportionality constant *α* $\approx$1.5 [8]. In Peyrusson et al., [8], *α* is estimated through measurements of cellular uptake of radiolabelled gepotidacin into human PMN.

##### S2.2 Modelling gentamicin and azithromycin pharmacokinetics.

The assumption of exponential decay of drug concentration implies instantaneous drug distribution and equilibration throughout relevant tissue [10]. However, data from studies involving azithromycin and gentamicin are inconsistent with this assumption [11, 12]. Azithromycin shows rapid entry into host cells and high intracellular accumulation followed by slow release into the extracellular environment [11, 13]. In the case of gentamicin, while it has been considered to principally kill extracellular NG due to its poor intracellular penetration [14], other studies have observed a slow increase in intracellular drug concentration that plateaus after about 3-4 days. [12, 15]. To capture these differences in drug distribution the antibiotic concentrations of gentamicin and azithromycin are captured through a two-compartment model where the extracellular ($C_{e}$) and intracellular ($C_{i}$) antibiotic concentrations are modelled separately according to Eq. S2 and S3:

$$\frac{dC_{e}}{dt}\text{ = - }\text{δ}C_{e}-k_{12}C_{e}+k_{21}C_{i}\frac{V_{i}}{V_{e}}. (S2)$$

$$\frac{dC_{i}}{dt}\text{ = }k_{12}C_{e}\frac{V_{e}}{V_{i}}-k_{21}C_{i}. (S3)$$

Here, $\text{δ}$ is the drug elimination rate constant, $k_{12}$and $k_{21}$ are respectively the rate constants of drug movement from and to the extracellular compartment, $V_{e}$ and $V_{i}$ are respectively the extracellular and intracellular volumes of distribution. The parameter values and sources for these are described in main text Table 1.

#### S3 Calibrating PK/PD parameters using susceptibility breakpoints.

When the model-derived susceptibility breakpoints are evaluated using the LHS samples derived from the Section 2.2 in the main text, in some cases the proportion of simulations that achieved clearance when the Hill function MIC parameter was set at ‘empirical breakpoints’ was lower than the expected 95% clearance rate. Here we define the term ‘empirical breakpoints’ to refer to susceptibility breakpoints published by the Clinical and Laboratory Standards Institute (CLSI) and the European Committee on Antimicrobial Susceptibility Testing (EUCAST) or in relation to new candidate drugs from relevant published studies. In order to align the model-derived breakpoints with empirical breakpoints, we then decided to calibrate the model to these empirical breakpoints and thereby refine the ranges of the parameters that are influential in determining the model-derived susceptibility breakpoints.

In this parameter refinement process, we first analyse the importance of each parameter's uncertainty in contributing to the variability of the model-derived susceptibility breakpoints (influential parameters) through partial rank correlation coefficients (Fig. S3). In these influential parameters, parameter ranges are analysed of the simulations that achieve empirical susceptibility breakpoints and these show skewed parameter distributions (Figs S4- S6). For gepotidacin, as there is no defined empirical susceptibility breakpoint, the model-derived breakpoint of 0.64mg/L using the point estimates is used as the cut-off for the calibration process. Then, in these influential parameters, the parameter ranges that consist of 90% of the simulations that meet empirical susceptibility breakpoints are selected as the refined range that is used for the subsequent model simulations. A comparison between the original parameter values (before model calibration) and the refined parameter ranges (after calibration) is shown in Table S2. Using these refined treatment parameter ranges, a new set of LHS samples are generated for the treatment parameters (PK/PD related) and are then combined with the LHS samples from the natural infection model as described in Section 2.2 in main text.

Table S2: Comparison of the parameter values prior to calibration and after calibration for the three antibiotics gepotidacin (GEP), gentamicin (GEN) and azithromycin (AZM).

| Drug | Parameter | Parameter range (prior to calibration) | Reference for parameter values prior to calibration | Parameter range (after calibration) |
| --- | --- | --- | --- | --- |
| GEP | $\varphi_{min}$ ${(h}^{-1})$ | -0.64, -0.43 | Fitting to time-kill data in Farrell, Sader (3) | -0.64, -0.46 |
|  | $k_{H}$ | 1.3 – 3.64 | Fitting to time-kill data in Farrell, Sader (3) | 1.78 – 3.64 |
| GEN | $V_{e}$ (L) | 0.28 – 1.29 | [16] | 0.6 – 1.29 |
|  | $k_{12}$ ${(h}^{-1})$ | 0.01 – 0.04 | [16] | 0.03 – 0.04 |
| AZM | $V_{e}$ (L) | 358 – 779 | [17] | 485 – 779 |
|  | $V_{i}$ (L) | 981 – 2577 | [17] | 981 – 1916 |
|  | $k_{12}$ ${(h}^{-1})$ | 0.07 – 0.18 | [17] | 0.1 – 0.18 |
|  | $k_{21}$ ${(h}^{-1})$ | 0.02 – 0.06 | [17] | 0.03 – 0.06 |

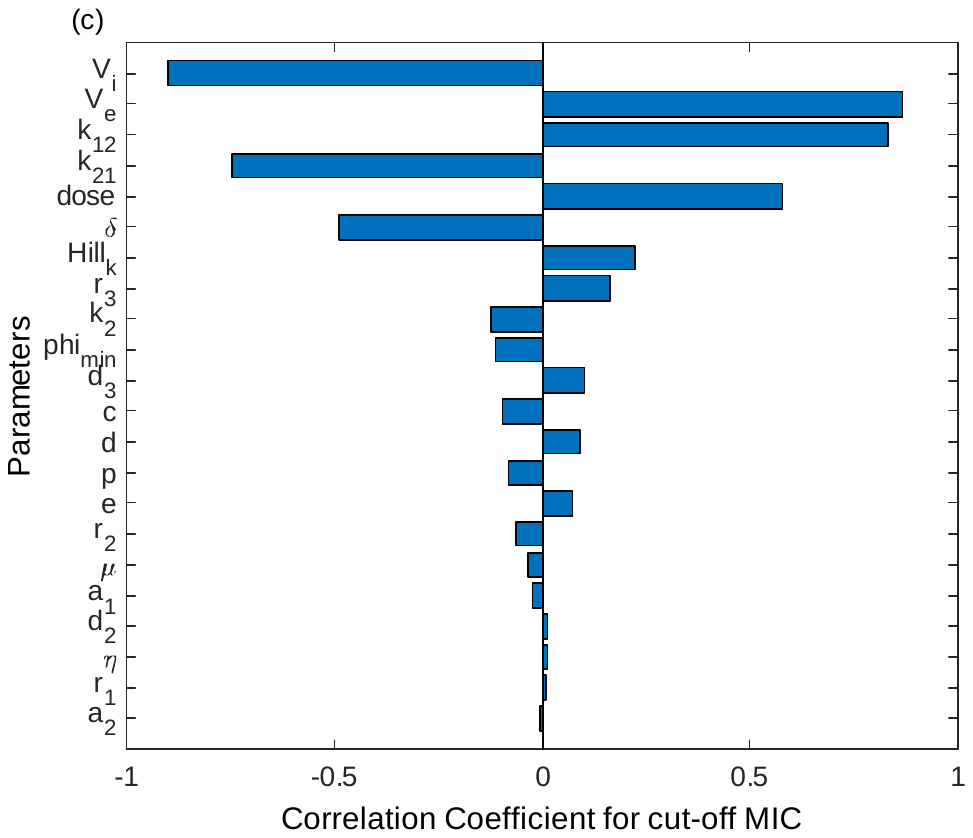

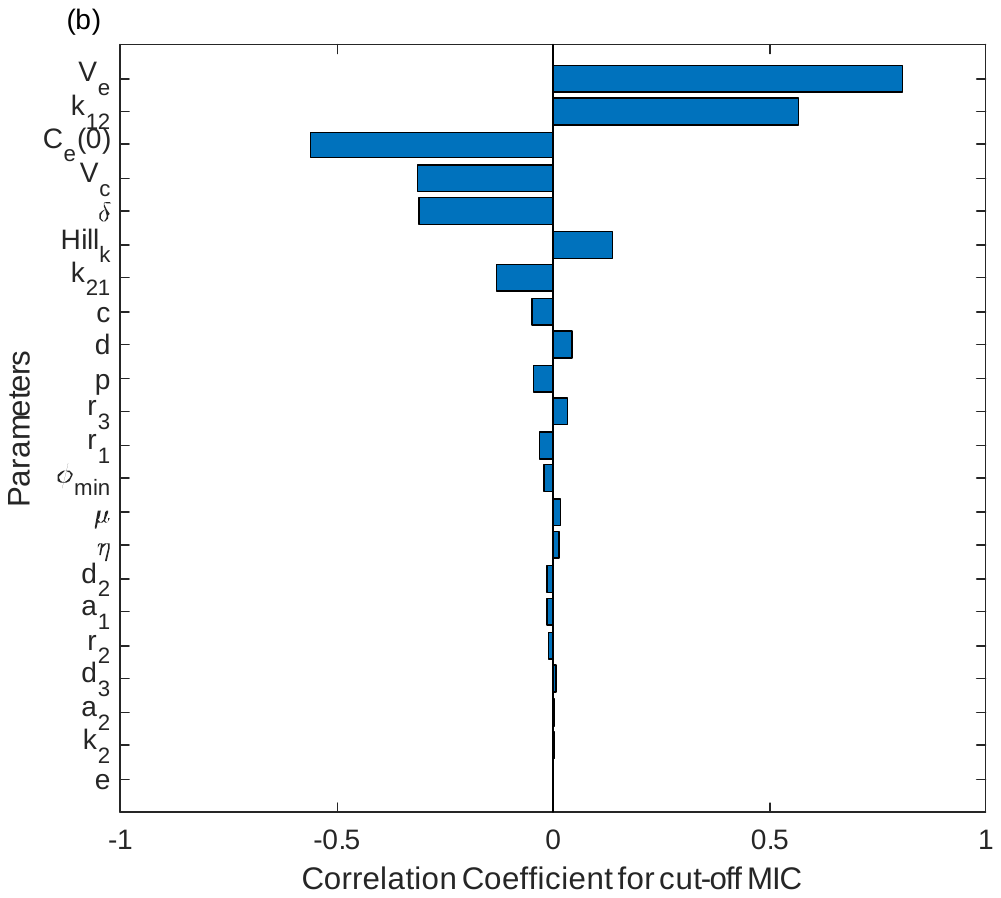

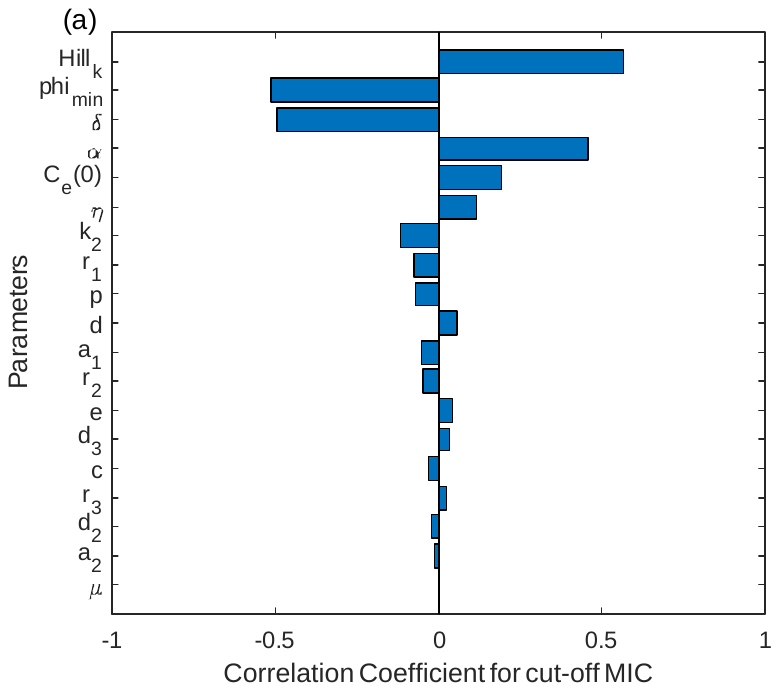

Figure S3: Tornado plots of partial rank correlation coefficients, indicating the importance of each parameter's uncertainty in contributing to the variability in the model-derived susceptibility breakpoint obtained from the simulations of LHS samples shown for (a) gepotidacin, (b) gentamicin and (c) azithromycin.

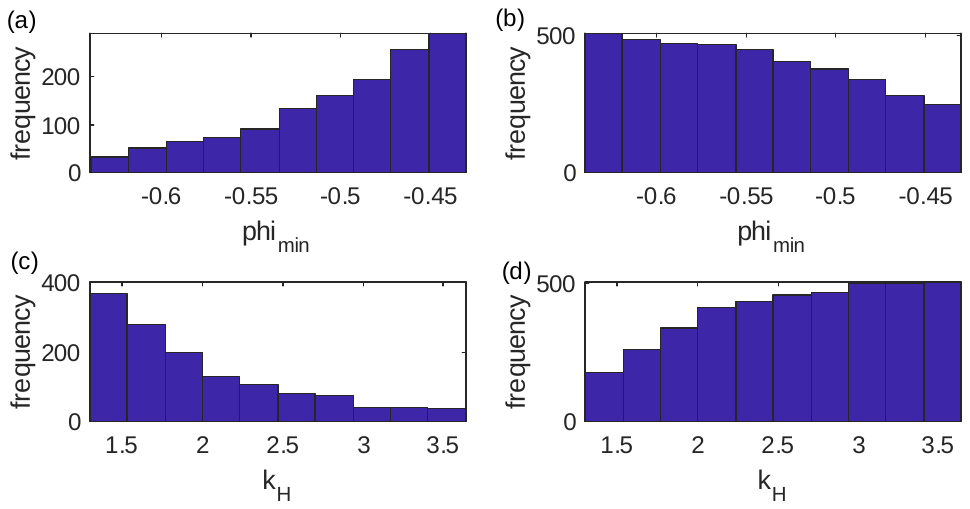

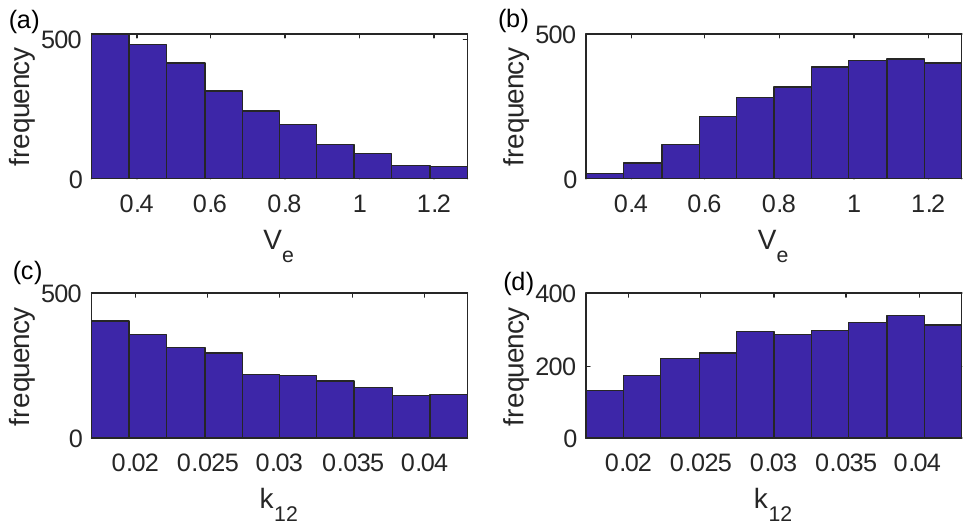
Figure S4: Parameter distributions of the simulations from LHS samples that resulted in a model-derived susceptibility breakpoint < 0.64mg/L (left column) and ≥ 0.64mg/L (right panel) shown for the parameters that were indicated as influential for the variability of gepotidacin susceptibility breakpoint ($\varphi_{min}, k_{H}$).

Figure S5: Parameter distributions of the simulations from the LHS samples that resulted in a model-derived susceptibility breakpoint < 4mg/L (left column) and ≥ 4mg/L (right panel) shown for the parameters that were indicated as influential for the variability of gentamicin susceptibility breakpoint level
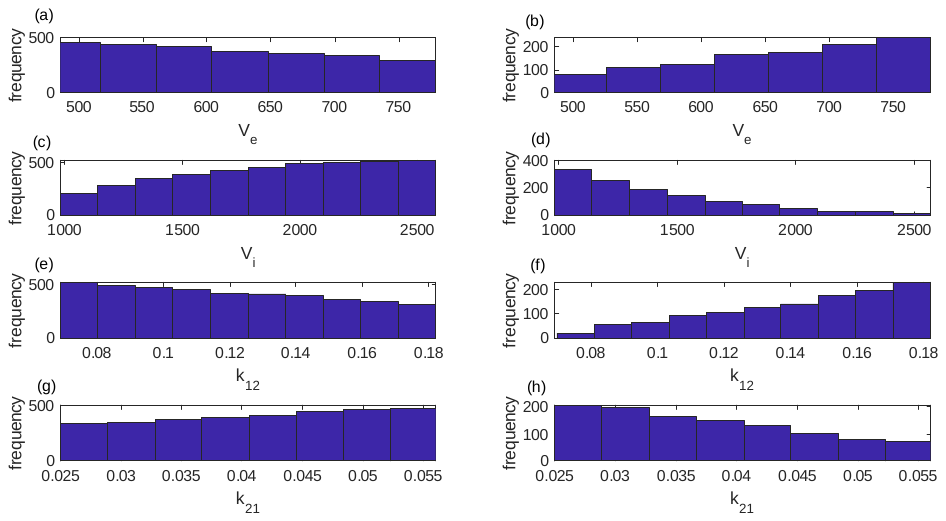
($V_{e}, k_{12}$).

Figure S6: Parameter distributions of the simulations from the LHS samples that resulted in a model-derived susceptibility breakpoint < 1mg/L (left column) and ≥ 1mg/L (right panel) shown for the parameters that were indicated as influential for the variability of azithromycin susceptibility breakpoint ($V_{e}, V_{i}, k_{12},k_{21}$).

#### S4 Model equations

Model equations when treatment is included to the four NG states (unattached NG (*B*), attached NG ($B_{a}$) to epithelial cells, NG internalised within epithelial cells ($B_{i}$) and NG surviving within PMN ($B_{s}$)) are described in this section. The innate immune response through PMN (*N*) is the considered immune response in the model. Treatment is included based on the drug pharmacokinetics as described in the main file. For parameter values and notations refer main text Table 1.

##### S4.1 Antibiotic concentration modelled as a one-compartment (gepotidacin)

$$\frac{dB}{\mathrm{dt}}=\left( 1-\frac{B+B_{a}}{k_{1}} \right)\left( r_{1} B+d_{3} B_{s}+eB_{i} \right)-d\frac{B N}{c N+B}-d_{2} B-a_{1} B\left( 1-\frac{B_{a}}{k_{1} a_{2}} \right)- \frac{\left( r_{1}-\varphi_{min} \right)\left( \frac{C_{e}}{MIC} \right)^{k_{H}}}{\left( \frac{C_{e}}{MIC} \right)^{k_{H}} - \frac{\varphi_{min}}{r_{1}}} B\left( 1-\frac{B+B_{a}}{k_{1}} \right)$$

$$\frac{dB_{a}}{\mathrm{dt}}=r_{1}B_{a}\left( 1-\frac{B+B_{a}}{k_{1}} \right)+a_{1} B\left( 1-\frac{B_{a}}{k_{1} a_{2}} \right)-d\frac{B_{a} N}{c N+B_{a}}-\eta B_{a}- \frac{\left( r_{1}-\varphi_{min} \right)\left( \frac{C_{e}}{MIC} \right)^{k_{H}}}{\left( \frac{C_{e}}{MIC} \right)^{k_{H}} - \frac{\varphi_{min}}{r_{1}}} B_{a}\left( 1-\frac{B+B_{a}}{k_{1}} \right)$$

$$\frac{dB_{i}}{\mathrm{dt}}=\left( 1-\frac{B_{i}}{k_{1}a_{2}} \right)\left( \eta B_{a}+r_{2}B_{i} \right)-eB_{i}- \frac{\left( r_{2}-\varphi_{min} \right)\left( \alpha\frac{C_{e}}{MIC} \right)^{k_{H}}}{\left( \alpha\frac{C_{e}}{MIC} \right)^{k_{H}} - \frac{\varphi_{min}}{r_{2}}} B_{i}\left( 1-\frac{B_{i}}{k_{1}a_{2}} \right)$$

$$\frac{dB_{s}}{\mathrm{dt}}=\left( 1-\frac{B_{s}}{{N k}_{2}} \right)\left( p d\frac{B N}{c N+B}+p d\frac{B_{a} N}{c N+B_{a}}+r_{3} B_{s} \right)-d_{3}B_{s}- \frac{\left( r_{3}-\varphi_{min} \right)\left( \alpha\frac{C_{e}}{MIC} \right)^{k_{H}}}{\left( \alpha\frac{C_{e}}{MIC} \right)^{k_{H}} - \frac{\varphi_{min}}{r_{3}}} B_{s}\left( 1-\frac{B_{s}}{{N k}_{2}} \right)$$

$$\frac{dN}{\mathrm{dt}}=\mu\left( N_{max}-N \right) \left( B+B_{a} \right)-d_{3} N$$

$$\frac{dC_{e}}{dt}\text{ = - }\text{δ}C_{e}$$

$C_{e}$ is the extracellular drug concentration.

##### S4.2 Model equations when antibiotic concentration modelled as a two-compartment model (azithromycin and gentamicin).

For these drugs extracellular ($C_{e}$) and intracellular ($C_{i}$) drug concentrations are modelled separately.

$$\frac{dB}{\mathrm{dt}}=\left( 1-\frac{B+B_{a}}{k_{1}} \right)\left( r_{1} B+d_{3} B_{s}+eB_{i} \right)-d\frac{B N}{c N+B}-d_{2} B-a_{1} B\left( 1-\frac{B_{a}}{k_{1} a_{2}} \right)- \frac{\left( r_{1}-\varphi_{min} \right)\left( \frac{C_{e}}{MIC} \right)^{k_{H}}}{\left( \frac{C_{e}}{MIC} \right)^{k_{H}} - \frac{\varphi_{min}}{r_{1}}} B\left( 1-\frac{B+B_{a}}{k_{1}} \right)$$

$$\frac{dB_{a}}{\mathrm{dt}}=r_{1}B_{a}\left( 1-\frac{B+B_{a}}{k_{1}} \right)+a_{1} B\left( 1-\frac{B_{a}}{k_{1} a_{2}} \right)-d\frac{B_{a} N}{c N+B_{a}}-\eta B_{a}- \frac{\left( r_{1}-\varphi_{min} \right)\left( \frac{C_{e}}{MIC} \right)^{k_{H}}}{\left( \frac{C_{e}}{MIC} \right)^{k_{H}} - \frac{\varphi_{min}}{r_{1}}} B_{a}\left( 1-\frac{B+B_{a}}{k_{1}} \right)$$

$$\frac{dB_{i}}{\mathrm{dt}}=\left( 1-\frac{B_{i}}{k_{1}a_{2}} \right)\left( \eta B_{a}+r_{2}B_{i} \right)-eB_{i}- \frac{\left( r_{2}-\varphi_{min} \right)\left( \frac{C_{i}}{MIC} \right)^{k_{H}}}{\left( \frac{C_{i}}{MIC} \right)^{k_{H}} - \frac{\varphi_{min}}{r_{2}}} B_{i}\left( 1-\frac{B_{i}}{k_{1}a_{2}} \right)$$

$$\frac{dB_{s}}{\mathrm{dt}}=\left( 1-\frac{B_{s}}{{N k}_{2}} \right)\left( p d\frac{B N}{c N+B}+p d\frac{B_{a} N}{c N+B_{a}}+r_{3} B_{s} \right)-d_{3}B_{s}- \frac{\left( r_{3}-\varphi_{min} \right)\left( \frac{C_{i}}{MIC} \right)^{k_{H}}}{\left( \frac{C_{i}}{MIC} \right)^{k_{H}} - \frac{\varphi_{min}}{r_{3}}} B_{s} \left( 1-\frac{B_{s}}{{N k}_{2}} \right)$$

$$\frac{dN}{\mathrm{dt}}=\mu\left( N_{max}-N \right) \left( B+B_{a} \right)-d_{3} N$$

$$\frac{dC_{e}}{dt}\text{ = }\text{- δ}C_{e}-k_{12}C_{e}+k_{21}C_{i}\frac{V_{i}}{V_{e}}$$

$$\frac{dC_{i}}{dt}\text{ = }k_{12}C_{e}\frac{V_{e}}{V_{i}}-k_{21}C_{i}$$

#### S5 Dual treatment

Drug interactions are commonly modelled using Loewe additivity [18] when their mechanisms of action or targets are similar, or Bliss independence [19] when these differ. In simple terms, Loewe additivity combines the effects of both drugs into a single drug of higher potency, while Bliss independence assumes independent multiplicative effects of the two drugs on bacterial survival. Macrolides (e.g. azithromycin) and aminoglycosides (e.g. gentamicin) disrupt bacterial protein synthesis by inhibiting ribosome functionality [20]. As gentamicin and azithromycin have similar targets and mechanisms of action [21, 22], we use the concept of Loewe additivity to model dual treatment effects, according to the method described in Dini et al., [23].

When drugs interact, they can show additive, synergistic or antagonistic effects. An additive effect is observed when the combined effect is similar to the effect obtained when the two drugs are administered individually. Additivity is taken as the reference level and if the observed combined effect is higher than the additive effect it is referred to as being synergistic while if the combined effect is lesser than the additive effect it is referred to as antagonistic. *In vitro* studies have tested drug synergism for treatment of NG strains and for the gentamicin + azithromycin combination no antagonistic or synergistic effects are observed [24-26]. Therefore, in the model, dual combination is tested under additive effect.

##### S5.1 Loewe additivity

To model under Loewe additivity, first, the concentration-drug effect relationship (obtained through $\frac{\left( \varphi_{max}-\varphi_{min} \right)\left( \frac{C}{MIC} \right)^{k_{H}}}{\left( \frac{C}{MIC} \right)^{k_{H}} - \frac{\varphi_{min}}{\varphi_{max}}}$) need to be analysed for gentamicin and azithromycin. Through this, the most dominant drug among the two can be identified.

Once the dominant drug is identified, the additive effect with the combination of azithromycin and gentamicin ($E_{ga}$) under Loewe additivity is given by Eq. S4 [23],

$$E_{ga}=\frac{\left( \varphi_{max,hp}-\varphi_{min,hp} \right)\left( \frac{C_{ag}}{{MIC}_{hp}} \right)^{k_{H,hp}}}{\left( \frac{C_{ag}}{{MIC}_{hp}} \right)^{k_{H,hp}} - \frac{\varphi_{min,hp}}{\varphi_{max,hp}}} (S4)$$

where $C_{ag}=C_{hp}+C_{eq}$ and $\varphi_{max,hp}$, $\varphi_{min,hp}$, $k_{H,hp}$ are the Hill function parameter values for the most potent drug out of gentamicin and azithromycin. $C_{hp}$ is the drug concentration of the more potent drug. $C_{eq}$ is the concentration of the more potent drug that is equally effective as the less potent drug at the concentration $C_{lp}$. This is obtained through,

$$C_{eq}=E^{-1}(E(C_{lp})) (S5)$$

where $E^{-1}$ is the inverse function of *E* given by,

$$E^{-1}\left( x \right)={MIC}_{hp} \left( \frac{\varphi_{min,hp} x}{\varphi_{max,hp} \left( x-\left( \varphi_{max,hp}-\varphi_{min,hp} \right) \right)} \right)^{\frac{1}{k_{H,hp}}} (S6)$$

and $E(C_{lp})$ is the Hill function effect obtained under the less potent drug at concentration $C_{lp}$.

The model equations under Loewe additivity are as follows for the zero-interaction (additive effect) case:

$$\frac{dB}{\mathrm{dt}}=\left( 1-\frac{B+B_{a}}{k_{1}} \right)\left( r_{1} B+d_{3} B_{s}+eB_{i} \right)-d\frac{B N}{c N+B}-d_{2} B-a_{1} B\left( 1-\frac{B_{a}}{k_{1} a_{2}} \right)-E_{ga} \left( 1-\frac{B+B_{a}}{k_{1}} \right)B$$

$$\frac{dB_{a}}{\mathrm{dt}}=r_{1}B_{a}\left( 1-\frac{B+B_{a}}{k_{1}} \right)+a_{1} B\left( 1-\frac{B_{a}}{k_{1} a_{2}} \right)-d\frac{B_{a} N}{c N+B_{a}}-\eta B_{a}- E_{ga} \left( 1-\frac{B+B_{a}}{k_{1}} \right)B_{a}$$

$$\frac{dB_{i}}{\mathrm{dt}}=\left( 1-\frac{B_{i}}{k_{1}a_{2}} \right)\left( \eta B_{a}+r_{2}B_{i} \right)-eB_{i}- E_{ga} \left( 1-\frac{B_{i}}{k_{1}a_{2}} \right)B_{i}$$

$$\frac{dB_{s}}{\mathrm{dt}}=\left( 1-\frac{B_{s}}{{N k}_{2}} \right)\left( p d\frac{B N}{c N+B}+p d\frac{B_{a} N}{c N+B_{a}}+r_{3} B_{s} \right)-d_{3}B_{s}- E_{ga} {\left( 1-\frac{B_{s}}{{N k}_{2}} \right)B}_{s}$$

$$\frac{dN}{\mathrm{dt}}=\mu\left( N_{max}-N \right) \left( B+B_{a} \right)-d_{3} N$$

#### S6 Different treatment strategies tested

In this study, we look at the effectiveness of several possible single and multiple dose treatment strategies by simulating the dosing regimens that are summarised in main text Tables 3 and 4. As a validation process, we assess treatment strategies that are tested in clinical trials as well as some possible novel strategies for gepotidacin monotreatment and gentamicin + azithromycin dual combination. For gepotidacin, we test single dose treatments of 1.5g and 3g as tested in clinical trials [27, 28] and higher single doses of 4.5g and 6g which have only been previously tested *in vitro* [29]. As novel gepotidacin treatment regimens, we also test multiple dose regimens including alternative spacing of doses. For the combination of gentamicin + azithromycin, we test the single dose options of 240mg gentamicin and 1g azithromycin [21] or 2g azithromycin [30] which have been previously tested in the respective clinical trials. We also consider several multiple dose regimens for the gentamicin + azithromycin combination that have not been previously tested in clinical trials. Here, for our multiple dose regimens using gentamicin, we choose a daily administration of 240mg which is the dose amount that has been tested in clinical trials [21, 30, 31], and limit the duration of dosing to 3 days basing on medical advice relating to toxicity concerns [32]. In addition, we conduct a limited analysis of patient non-adherence, whereby the second dose is delayed by uniformly distributed time between 0 and 8 hours, with any subsequent doses then taken at the scheduled delay from the previous dose.

#### S7 Gepotidacin monotreatment

##### S7.1 Association between model-derived breakpoint MIC and drug dose.

The following system given in Eq. S7 and S8 are non-dimensionalised to analyse whether the model-derived breakpoint MIC increases linearly with drug dose.

$$\frac{dB}{\mathrm{dt}}=\left( 1-\frac{B}{k_{1}} \right)r_{1} B - \frac{\left( r_{1}-\varphi_{min} \right)\left( \frac{C_{e}}{MIC} \right)^{k_{H}}}{\left( \frac{C_{e}}{MIC} \right)^{k_{H}} - \frac{\varphi_{min}}{r_{1}}} B\left( 1-\frac{B}{k_{1}} \right) (S7)$$

$$\frac{dC_{e}}{dt}\text{ = - }\text{δ}C_{e} (S8)$$

$B\left( 0 \right)=B_{0}$ and $C_{e}(0)= C_{0}$

Let $B=B^{*} \hat{B}$, $C_{e}={C_{e}}^{*} \hat{C_{e}}$ and *t*$=t^{*} \hat{t}$ such that the new variables $B^{*}$, $C^{*}$ and $t^{*}$ are non- dimensionalised versions of the original variables $B$, $C_{e}$ and *t* respectively. The values of the constants $\hat{B}$, $\hat{C_{e}}$ and $\hat{t}$ are determined later.

Writing the system in terms of the non- dimensionalised variables.

$$\frac{dB^{*} \hat{B}}{dt^{*} \hat{t}}=\left( 1-\frac{B^{*} \hat{B}}{k_{1}} \right)r_{1} B^{*} \hat{B} - \frac{\left( r_{1}-\varphi_{min} \right)\left( \frac{{C_{e}}^{*} \hat{C_{e}}}{MIC} \right)^{k_{H}}}{\left( \frac{{C_{e}}^{*} \hat{C_{e}}}{MIC} \right)^{k_{H}} - \frac{\varphi_{min}}{r_{1}}} B^{*} \hat{B}\left( 1-\frac{B^{*} \hat{B}}{k_{1}} \right) (S9)$$

$$\frac{d{C_{e}}^{*} \hat{C_{e}}}{dt^{*} \hat{t}}\text{ = - }\text{δ}{C_{e}}^{*} \hat{C_{e}} (S10)$$

Simplifying Eq. S9 and S10 yields,

$$\frac{dB^{*}}{dt^{*}}= \hat{t}\left( 1-\frac{B^{*} \hat{B}}{k_{1}} \right)r_{1} B^{*} - \hat{t} \frac{\left( r_{1}-\varphi_{min} \right)\left( \frac{{C_{e}}^{*} \hat{C_{e}}}{MIC} \right)^{k_{H}}}{\left( \frac{{C_{e}}^{*} \hat{C_{e}}}{MIC} \right)^{k_{H}} - \frac{\varphi_{min}}{r_{1}}} B^{*} \left( 1-\frac{B^{*} \hat{B}}{k_{1}} \right) (S11)$$

$$\frac{d{C_{e}}^{*}}{dt^{*}}\text{ = }-\hat{t}\text{ }\text{δ}{C_{e}}^{*} \left( S12 \right)$$

Choose $\hat{B}$, $\hat{C_{e}}$ and $\hat{t}$ such that,

$\hat{B}=k_{1}$, $\hat{C_{e}}=MIC$ and $\hat{t}=\frac{1}{\delta}$

Then Eq. S11 and S12 simplifies as,

$$\frac{dB^{*}}{dt^{*}}=\left( 1-B^{*} \right)\frac{r_{1}}{\text{δ}}B^{*}-\frac{1}{\text{δ}}\left[ \frac{\left( r_{1}-\varphi_{min} \right)\left( {C_{e}}^{*} \right)^{k_{H}}}{\left( {C_{e}}^{*} \right)^{k_{H}} - \frac{\varphi_{min}}{r_{1}}} \right]B^{*}\left( 1-B^{*} \right) \left( S13 \right)$$

$$\frac{d{C_{e}}^{*}}{dt^{*}}\text{ = }-{C_{e}}^{*} \left( S14 \right)$$

$B^{*}\left( 0 \right)=\frac{B_{0}}{k_{1}}$ and

${C_{e}}^{*}\left( 0 \right)=\frac{C_{0}}{MIC}$

##### S7.2 PK indices to explain effectiveness of different treatment strategies

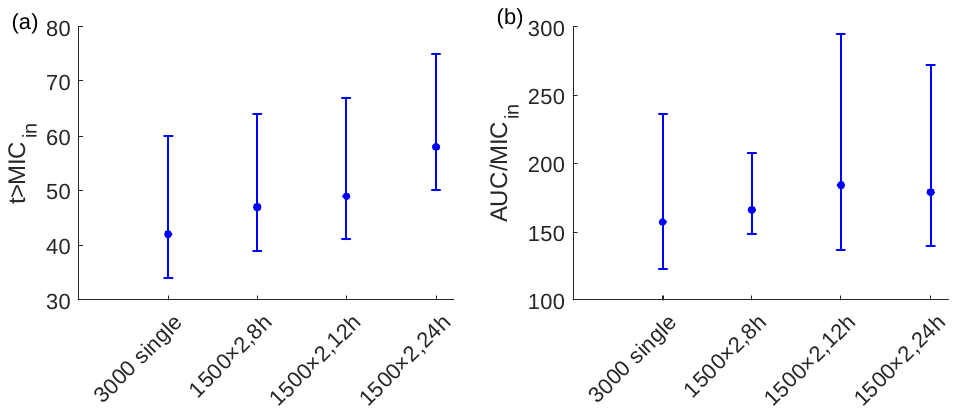
The higher effectiveness of multiple dose strategies than single dose regimens is explored using the PK indices, $t_{\mathrm{MIC}_{\mathrm{in}}}$, and $AUC/\mathrm{MIC}_{\mathrm{in}}$ (Fig.S7).

Figure S7: PK/PD indices evaluated for the 3000mg single dose and multiple dose strategies using 1500mg multiples (with a total accumulation of 3000mg) with dosing intervals of 8, 12 and 24h. (a) time above the MIC $(t_{\mathrm{MIC}_{\mathrm{in}}}$) and (b) area under the total drug concentration $(AUC/\mathrm{MIC}_{\mathrm{in}}$) calculated using the intracellular concentration. The data points represent the median and the bars represent the 95% range of the PK indices evaluated for the 5402 concentration profiles of the LHS samples for MIC for gepotidacin of 1mg/L.

##### S7.3 Threshold gepotidacin concentration required for treatment success

For multiple dose regimens we calculate the time above the MIC using our default method of total time the drug concentration remains above the MIC. We also calculate time above the MIC according to three alternative methods, 1) Total time above the MIC – Total time below the MIC (up till the last threshold crossing); 2) maximal continuous-time above the MIC (the longest period in which the drug concentration is always above the MIC); and 3) time above the MIC without considering the periods in which it is below the MIC (last time point the concentration is above MIC – first time point the concentration is above MIC). A comparison of these four index calculations is shown in Fig. S8.

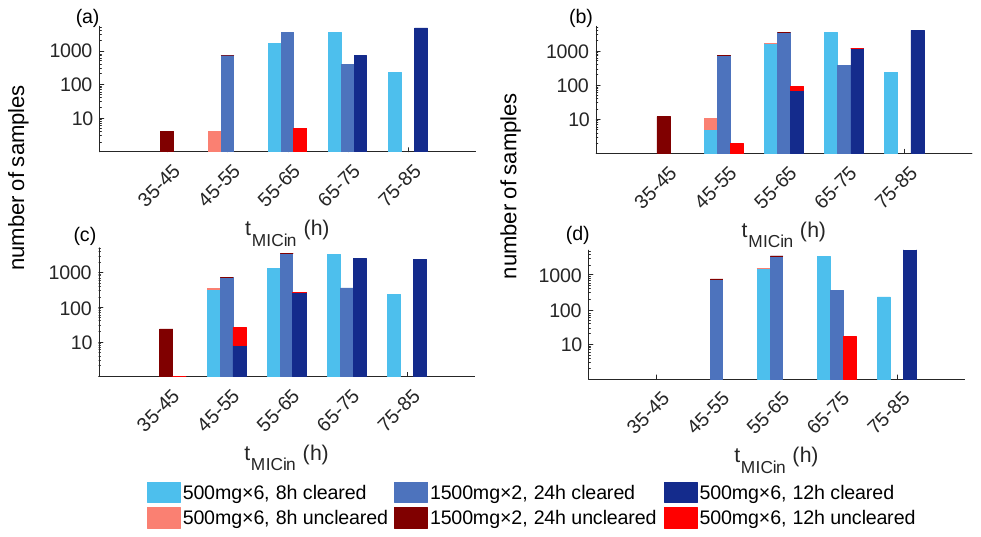

Figure S8: Comparison of $t_{\mathrm{MIC}_{\mathrm{in}}}$ calculations to differentiate treatment success and failure using intracellular drug concentration. (a) Total time above the MIC, removing periods where it is below; (b) total time above the MIC – total time below the MIC (up till the last threshold crossing); (c) maximal continuous-time above the MIC (the longest period in which the drug concentration is always above the MIC); and (d) time above the MIC without considering the periods in which it is below the MIC (last time point the concentration is above MIC – first time point the concentration is above MIC).

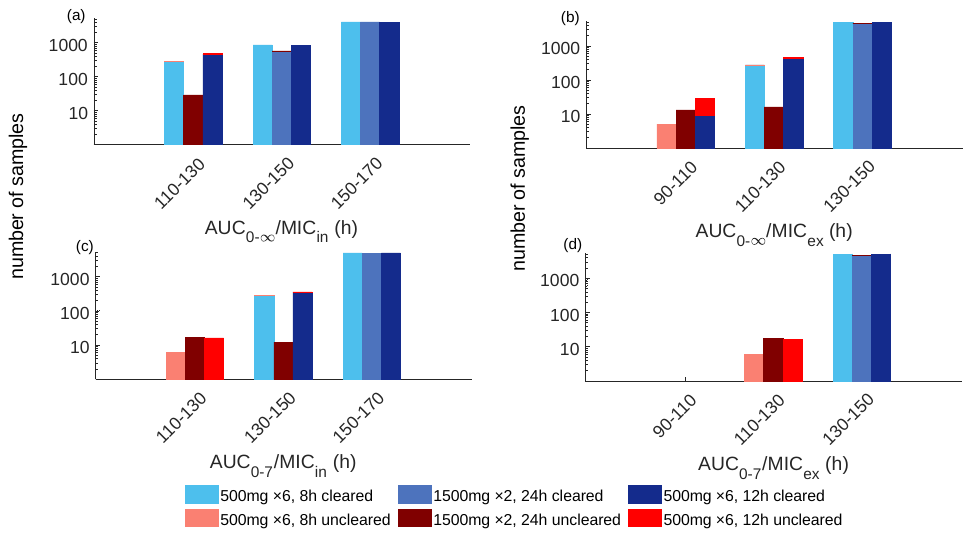
The area under the curve above the MIC (removing the area below the MIC from the total area under the curve) and AUC over a fixed time period of 7 days ($\mathrm{AUC}_{0-7}$/MIC) are calculated as alternative indices and the results are shown in Fig. S9.

Figure S9: The area under the curve above the MIC (removing the area below the MIC from the total area under the curve) (a, b) and AUC over a fixed time period of 7 days ($\mathrm{AUC}_{0-7}$/MIC) (c, d) calculated as alternative indices using intracellular drug concentration (first column) and (b) extracellular drug concentration (second column).

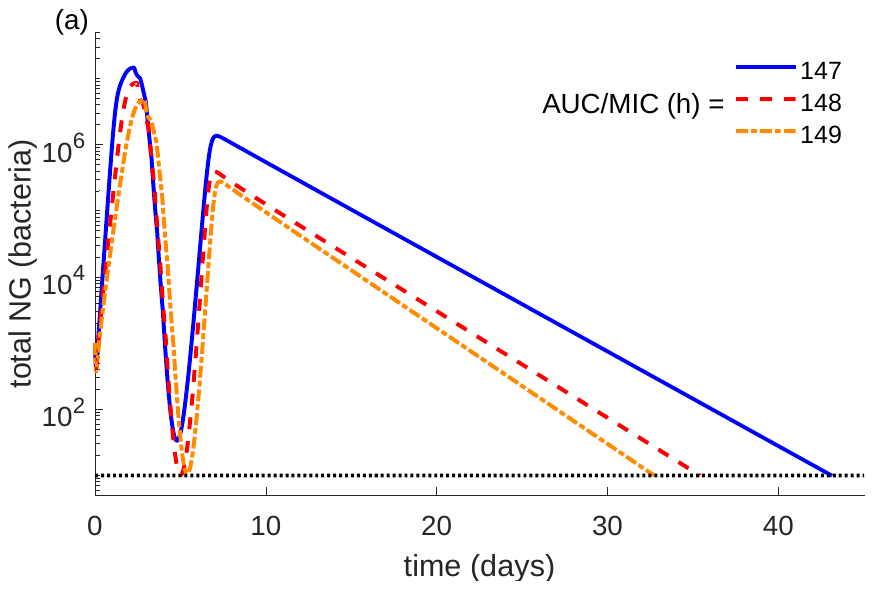
The behaviour of the bacterial load of the LHS samples that achieve $\mathrm{AUC}_{\mathrm{in}}/MIC$ in the range of 147-150h but fail to clear the infection is shown in Fig. S10.

Figure S10: Change in the total bacterial load over time of the samples that do not clear infection and have PK index $AUC/\mathrm{MIC}_{\mathrm{in}}$*<*150h*.* The shown bacterial load curves are associated with drug concentration levels that achieve $AUC/\mathrm{MIC}_{\mathrm{in}}$ in the range of 147-150h. Only three instances are shown for better visualisation.

##### S7.4 Testing higher doses of gepotidacin monotreatment

The *in vitro* study by VanScoy et al., [29] tested higher doses of 4.5 and 6g gepotidacin doses which have not been tested in clinical trials. The effectiveness of these higher doses is summarised in Table S3.

Table S3: Percentage of LHS samples (out of 5402) that clear the infection in ≤7 days when using single and multiple dose gepotidacin treatment strategies that accumulate to a 4.5 or 6g gepotidacin dose.

| Strategy | MIC (mg/L) | | | |
| --- | --- | --- | --- | --- |
|  | 1 | 1.5 | 2 | 2.5 |
| 4500mg single dose | 99.96 | 95.41 | 90.45 | 46.65 |
| 6000mg single dose | 100.00 | 99.74 | 95.41 | 81.89 |
| 1500mg ×3, 8h apart | 100.00 | 98.93 | 89.14 | 42.21 |
| 1500mg ×3, 12h apart | 100.00 | 99.24 | 90.76 | 67.42 |
| 1500mg ×3, 24h apart | 100.00 | 99.70 | 93.67 | 71.71 |
| 3000mg ×2, 8h apart | 100.00 | 99.81 | 96.87 | 87.23 |
| 3000mg ×2, 12h apart | 100.00 | 99.93 | 98.52 | 92.02 |
| 3000mg ×2, 24h apart | 100.00 | 99.98 | 99.35 | 93.29 |
| 2000mg×3, 8h apart | 100.00 | 99.10 | 98.93 | 87.75 |
| 2000mg×3, 24h apart | 100.00 | 99.84 | 99.24 | 91.13 |
| 4500mg on the first day and 1500mg on the second day | 100.00 | 99.98 | 98.93 | 92.65 |

#### S8 Gentamicin treatment.

##### S8.1 Impact on patient non-compliance on multiple-dose strategies of gentamicin

We analyse the impact of patient non-compliance for the extended gentamicin dosing strategy of 240mg × 3 doses, given 24h apart in combination with a 2g single dose of azithromycin. It is assumed the second dose will not be taken at the appropriate dosing interval. Treatment efficacy is analysed when 15%, 25% 50%, 75% and 100% of the LHS samples are assumed to be subject to non-adherence (Table S4).

Table S4: Treatment effectiveness with non-compliance for the dual treatment combination 240mg × 3 gentamicin, given 24h apart in combination with 2g azithromycin.

| Compliance level | (Gentamicin/ azithromycin) MIC (mg/L) | | | | | |
| --- | --- | --- | --- | --- | --- | --- |
|  | 4/0.5 | 4/1 | 8/0.5 | 8/1 | 16/0.5 | 16/1 |
| Full compliance | 100.00 | 99.91 | 99.96 | 98.96 | 99.76 | 95.45 |
| 15% non-compliance | 100.00 | 99.91 | 99.89 | 98.78 | 99.44 | 95.15 |
| 25% non-compliance | 100.00 | 99.91 | 99.91 | 98.78 | 99.44 | 94.75 |
| 50% non-compliance | 100.00 | 99.89 | 99.89 | 98.80 | 99.44 | 94.30 |
| 75% non-compliance | 100.00 | 99.89 | 99.87 | 98.76 | 99.43 | 94.13 |
| 100% non-compliance | 100.00 | 99.85 | 99.85 | 98.74 | 99.41 | 94.13 |

32.	Queensland Health. Aminoglycoside Dosing in Adults 2018.
